# Spatial structure and demographic decoupling of chikungunya transmission and severity in Brazil, 2015 to 2025

**DOI:** 10.64898/2026.06.26.26356655

**Authors:** Quanqi Zhang, Fabricio Souza Campos, Filipe Vieira Santos de Abreu, William M. de Souza, Siyu Chen, Ana I. Bento

## Abstract

Brazil bears one of the largest chikungunya burdens in the Americas (more than 1·2 million confirmed cases since 2014), and the 2024–25 wave expanded further south than any prior outbreak. IXCHIQ entered *Sistema Único de Saúde* (SUS) deployment in February 2026 despite an August 2025 US FDA biologics license suspension over serious adverse events in adults aged 60 years and older; regulatory authorities in the EU (EMA), Brazil (ANVISA), and Canada maintained licensing with revised prescribing guidance requiring individual benefit–risk assessment. Evidence to guide rollout targeting is lacking.

We analyzed 1,235,424 confirmed chikungunya cases reported to *Sistema de Informação de Agravos de Notificação* SINAN (2015–2025) across 5,570 municipalities. A Bayesian hierarchical spatiotemporal model quantified spatial structure and persistence of transmission across municipalities, controlling for national arboviral co-circulation trends. Disease progression was assessed with Cox models stratified by age and sex. Municipal 2024–25 incidence rate ratios and proportions aged 65 years or older (tertiles) defined transmission-control, clinical-preparedness, and combined-priority municipalities.

Transmission epicenters shifted from the Northeast (2016–17) to the Central-West (2024–25; peak municipality IRRs >2·7× the national median); clustering and seasonality persisted. Cases concentrated among adults aged 25 to 55, while post-hospitalization mortality rose steeply with age (HR 10·57, 95% CI 7·64–14·62 for ages 80 years and older versus adults aged 20 to 29). Males had faster progression to hospitalization (HR 1·26) and death (HR 1·76, onset to death) despite fewer notifications. The Central-West led transmission yet had lower case fatality among hospitalized cases (3·54%, 81 deaths out of 2,285 hospitalized) than the Southeast (4·95%, 163 deaths out of 3,291 hospitalized), reflecting demographic rather than purely clinical differences between regions. A municipality-level allocation framework classified 832 municipalities as transmission-control priority (high recent transmission, younger population; predominantly Central-West), 832 as clinical-preparedness priority (lower transmission, older population; predominantly Southeast and South), and 461 as combined-priority (high on both dimensions; predominantly Northeast and Southeast).

## Introduction

Brazil bears one of the largest national chikungunya burdens in the Americas, with over one million laboratory-confirmed cases since 2014,^1^ and the 2024–25 epidemic wave expanded further south than any prior outbreak. Rio Grande do Sul reported its first chikungunya death in April 2025,^2^ marking a geographic front advancing into populations with lower prior exposure. Against this backdrop, Brazil approved the first chikungunya vaccine (IXCHIQ, Valneva, live attenuated) through ANVISA (*Agência Nacional de Vigilância Sanitária*) in April 2025 and began pilot deployment through the *Sistema Único de Saúde* (SUS) in February 2026 ^3–5^. The rollout proceeds despite an August 2025 US FDA safety action focused on adults aged 60 years and older after reports of serious adverse events, including encephalitis and vaccine-strain chikungunya-like illness,^3–5^ against rising chikungunya activity across the Americas in early 2026.^6,7^ The safety signal falls precisely on the demographic group bearing the greatest risk of severe disease, making evidence on where transmission is most intense and where severe outcomes concentrate not a background question but the central operational challenge for the ongoing rollout.

Where does chikungunya transmission concentrate geographically, and why does the demographic profile of those who contract it differ so sharply from those who die of it? Because the disease is transmitted primarily by *Aedes aegypti*, an urban vector whose abundance scales with human population density, a priori expectation favors structured geographic clustering over independent local dynamics.^8^ Previous nationwide work described recurrent regional outbreaks through 2022, phylogeographic analysis identified the Northeast as a recurrent source of interregional viral dispersal, and sub-municipal analyses in Salvador documented strong intraurban clustering, but no national study had extended through the 2024–25 Central-West expansion, tested independence from Zika co-circulation, or quantified persistence across epidemic cycles.^9–11^ On the severity side, national cohort analyses have shown that hospitalization, in-hospital mortality, and years of life lost concentrate in older adults, but none have linked those severity gradients to geographic transmission structure.^12–14^

A recent national modeling study estimated force of infection at Brazil’s state level.^12^ We test that homogeneity assumption empirically, quantify spatial structure, and integrate it with demographic severity gradients to produce a municipality-level allocation framework for the ongoing rollout. A concurrent nationwide SINAN cohort documented age, sex, and comorbidity gradients in hospitalization, mortality, and years of life lost through 2024.^14^ We ask where those high-risk individuals concentrate relative to transmission intensity. If transmission and severity were geographically co-located, an age-targeted rule would suffice; their decoupling motivates the dual-axis framework. Answering both is urgent because the SUS rollout began in February 2026 without this evidence.

## Methods

### Data sources and study population

We conducted a nationwide retrospective observational study of 1,235,424 confirmed chikungunya cases reported to SINAN (*Sistema de Informação de Agravos de Notificação*) via DATASUS^15^ from January 1, 2015, to December 31, 2025, across all 5,570 Brazilian municipalities. Confirmed cases encompass laboratory-confirmed (n = 462,849; RT-PCR, viral isolation, or serology), clinically confirmed (n = 752,444; compatible symptoms and epidemiological linkage), and cases under investigation (n = 19,892); suspected and discarded cases were excluded. Analyses were restricted to laboratory-confirmed cases per SINAN classification criteria; suspected or discarded notifications were excluded.^12,16^ Individuals with missing race or ethnicity (23·2%) were excluded from race-stratified analyses; full data and denominator details are in the appendix.^15,17,19^

### Statistical analyses

National and regional temporal trends were summarized using weekly aggregated case counts and population-standardized incidence rates. Temporal periodicity was characterized using a Morlet wavelet transform of monthly national case counts, assessed against an autoregressive order-one red-noise background.^20^

Municipality-month counts were modeled using a Bayesian hierarchical spatiotemporal Poisson model with a Besag–York–Mollié 2 (BYM2) spatial effect,^21^ a second-order random walk temporal trend, an unstructured space–time interaction, and standardized national monthly Zika incidence, selected over dengue because its temporally distinct epidemic peak provides a cleaner test of whether chikungunya spatial gradients mirror broad arboviral surveillance pressure, whereas dengue’s long-standing endemicity would introduce collinearity, as a covariate, fitted with INLA (appendix). As a sensitivity analysis, the model was refitted with a negative binomial likelihood.^22,24^ Global Moran’s I and Local Indicators of Spatial Association (LISA) were computed on the posterior space–time interaction term to test for residual spatial clustering at each monthly time step (appendix).

Disease progression was analyzed using Kaplan-Meier methods and Cox proportional hazards models for three sequential intervals: symptom onset to hospitalization, hospitalization to death, and symptom onset to death, stratified by age group, sex, and race or ethnicity.^25,26^ Cox models were fitted univariably to characterize population-level demographic gradients; a mutually adjusted sensitivity analysis is reported in the appendix. The proportional hazards assumption was evaluated using Schoenfeld residuals.

All analyses used R (version 4·2 or later) with the sf, spdep, INLA, survival, survminer, and WaveletComp packages; full methodological detail is provided in the appendix. Reporting follows STROBE recommendations for observational studies and RECORD guidance for studies using routinely collected health data, where applicable.

### Data sharing

This study used anonymized, publicly available surveillance data; ethical approval was not required under Brazilian regulations. All data and analysis code are available at https://github.com/hemaanizg/Chikungunya-Spatiotemporal-Analysis.

## Results

Between January 2015 and December 2025, approximately 1·2 million confirmed chikungunya cases were reported to SINAN across all 5,570 Brazilian municipalities. National incidence showed two epidemic peaks in 2016-17 and 2023-24, each well above the long-term weekly average and separated by extended interepidemic periods (Figure 1A). Wavelet analysis identified a dominant annual cycle (peak period ≈1 year; p<0·01 against a red-noise background), amplified during both waves, with no sustained multi-year periodicity (Figure S2). The demographic and racial or ethnic profiles of reported cases, concentrated among adults aged 25–55 years with a female majority and predominance of Pardo individuals, were stable across epidemic years (Figure 1B–D; Figure S1). Because laboratory confirmation rates vary substantially across municipalities, incidence rate ratios throughout this analysis reflect gradients in confirmed disease burden and should be interpreted as planning heuristics rather than measures of true infection incidence.

**Figure 1:**
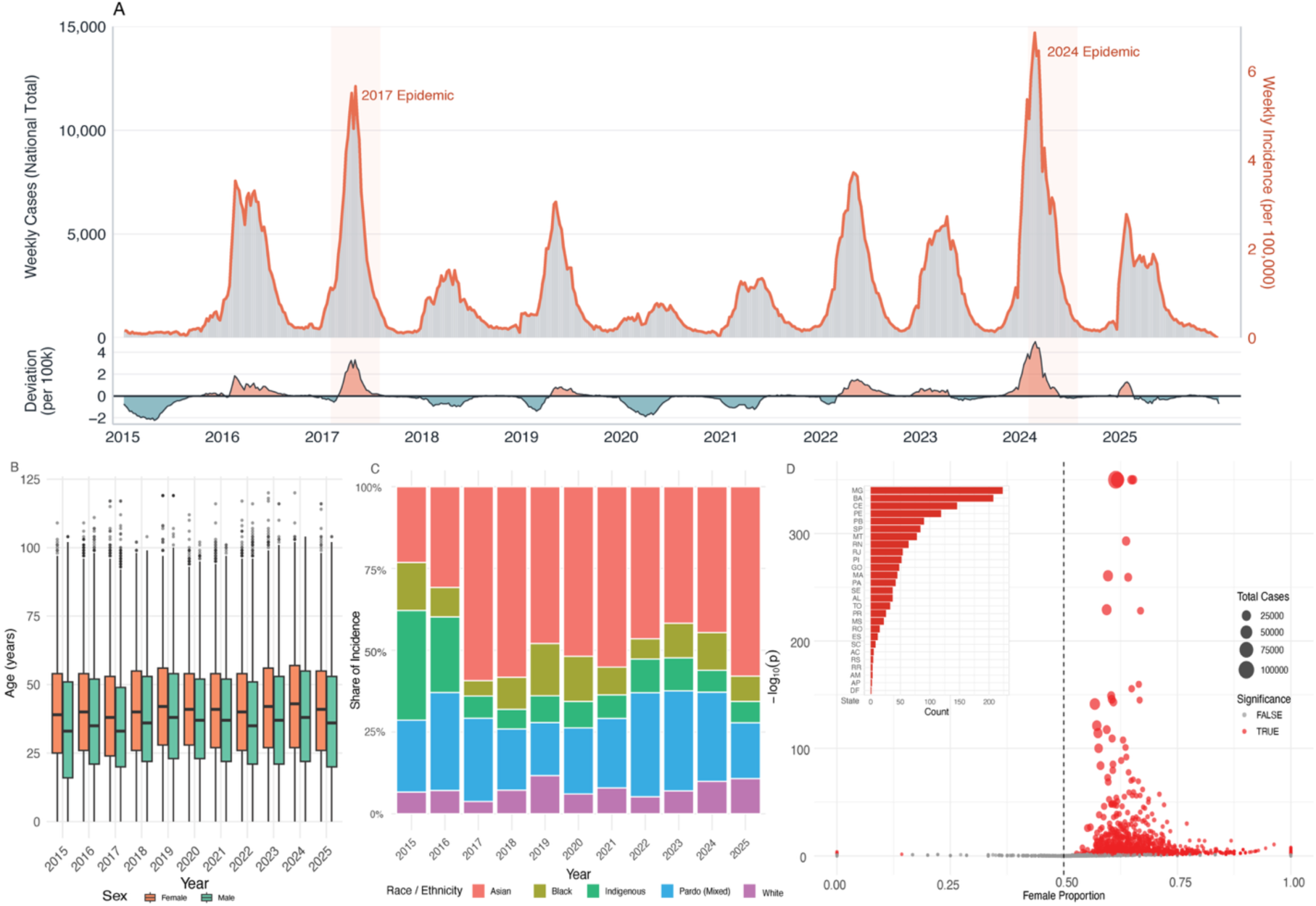
National chikungunya incidence and demographic profile of reported cases, Brazil, 2015 to 2025. (A) Weekly national incidence of laboratory-confirmed chikungunya cases per 100,000 population reported to SINAN, January 2015 to December 2025. The dashed horizontal line indicates the long-term weekly mean. Two major epidemic waves are visible, peaking in 2016 to 2017 and 2023 to 2024, separated by extended interepidemic troughs. (B) Annual age distribution of reported cases, shown as boxplots across municipalities. Cases were consistently concentrated among adults aged 25 to 55 years, and this profile did not change appreciably between epidemic and interepidemic years. (C) Annual incidence by race or ethnicity category, shown as population-standardized rates. (D) Municipality-level sex ratio of reported cases. Each point represents one municipality; the x-axis shows the absolute difference between female and male case counts, and the y-axis shows the statistical significance of a binomial test. Municipalities above the significance threshold with a positive difference indicate female excess in notifications, consistent with higher care-seeking and laboratory confirmation probabilities among females.

The geographic center of transmission shifted across epidemic waves, but the underlying spatial structure persisted throughout the surveillance period (Figure 2). In 2015 to 2017, the Northeast accounted for the largest deviations above the national municipality-median incidence (log IRR 0·3 to 0·6, corresponding to peak incidence of 225 to 242 cases per 100,000; Figure S3A). By 2024-2025, the Central-West had become the dominant high-transmission region (log IRR greater than 1·0, corresponding to more than a two-fold excess over the national median and peak incidence of 194 to 327 cases per 100,000), representing the largest relative shift in model-estimated transmission intensity across macro-regions during the surveillance era.

**Figure 2:**
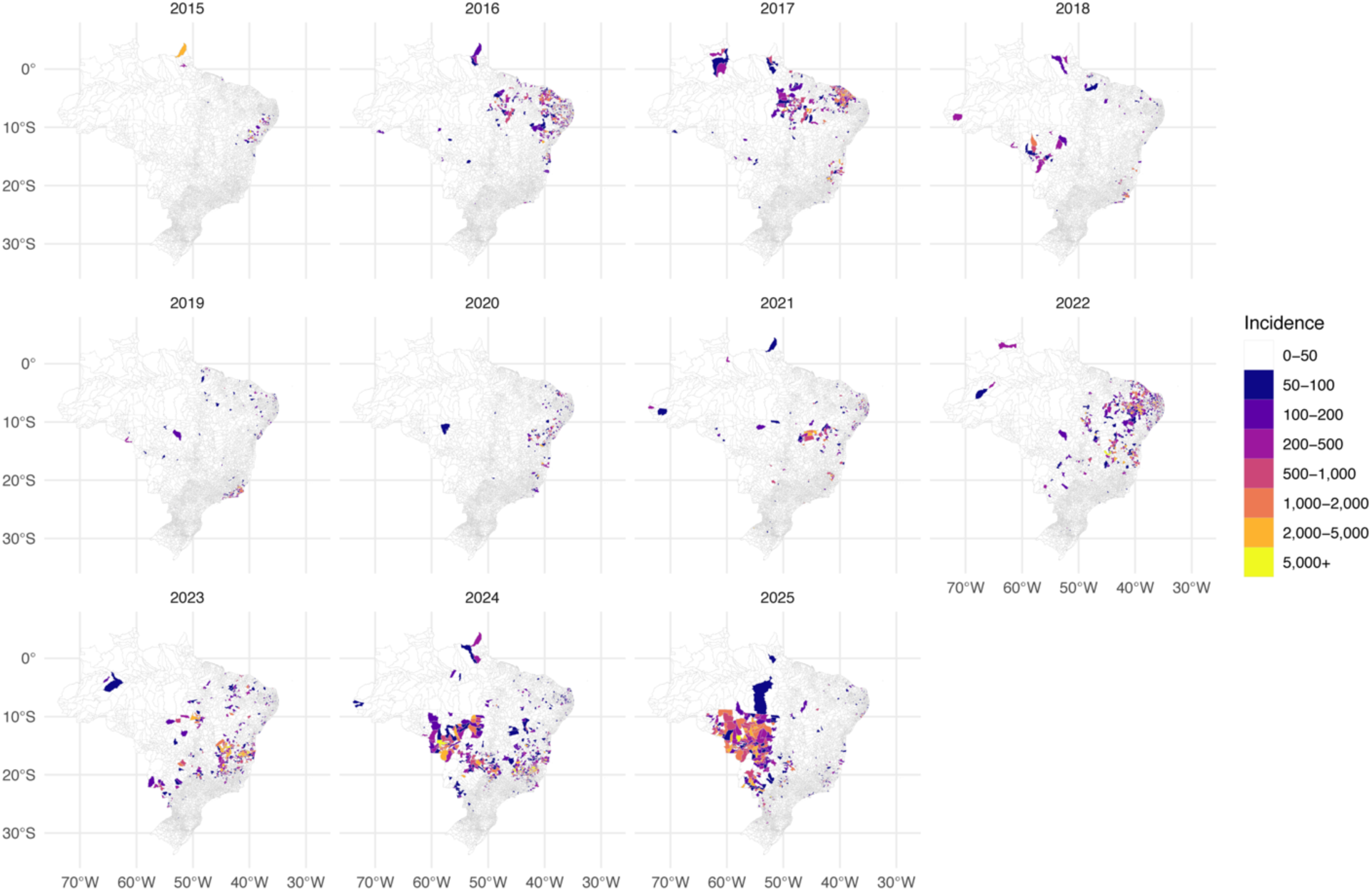
Geographic shift in chikungunya transmission intensity across Brazilian macro-regions, 2015 to 2025. Maps show municipality-level estimated incidence rate ratios (IRR) relative to the national municipality-median incidence at each time point, derived from the Bayesian spatiotemporal model. Warm colors indicate municipalities with incidence above the national median; cool colors indicate municipalities below. All panels share a common color scale to permit direct comparison across time periods. In 2015 to 2017, the highest transmission was concentrated in the Northeast (log IRR 0·3 to 0·6). By 2024 to 2025, the Central-West had emerged as the dominant high-transmission region (log IRR greater than 1·0), representing the largest geographic expansion of chikungunya in Brazil during the surveillance era.

The Bayesian hierarchical spatiotemporal model confirmed the strength of this spatial structure and its independence from Zika co-circulation. Approximately 70% of the spatial random-effect variance was attributable to structured geographic clustering among neighboring municipalities (*φ=* 0·70, 95% CrI 0·65 to 0·76; Figure 3B). The structured spatial random effect ranged from −3·66 to +4·46 on the log scale, corresponding to more than an 80-fold difference in baseline incidence between the most and least affected municipalities. Global Moran’s I of the space-time interaction was positive and significant in all 132 monthly time steps (range 0·047 to 0·503; median 0·215; p <0·05 in every month; Figure 3A). The wide range of individual space-time deviations (−4·73 to +10·04 on the log scale) reflects genuine municipality-specific epidemic timing rather than model misspecification, as confirmed by the persistent positive Moran’s I across all time steps. The temporal random effect ranged from −2·21 to +2·26 on the log scale (Figure S3C), corresponding to more than a ninefold difference between epidemic peaks and interepidemic troughs. National Zika co-circulation, incorporated as a time-varying covariate to test whether CHIKV spatial structure could be explained by shared arboviral surveillance pressure, showed no independent association with chikungunya incidence after conditioning on spatial and temporal random effects (exp(β) 1·12, 95% CrI 0·94 to 1·33), indicating that the chikungunya spatial structure is not explained by national-level Zika trends, though municipality-level co-circulation remains incompletely captured. A negative binomial sensitivity analysis yielded highly consistent spatial-structure estimates (φ = 0·693, 95% CrI 0·641 to 0·744; Table S4).

**Figure 3:**
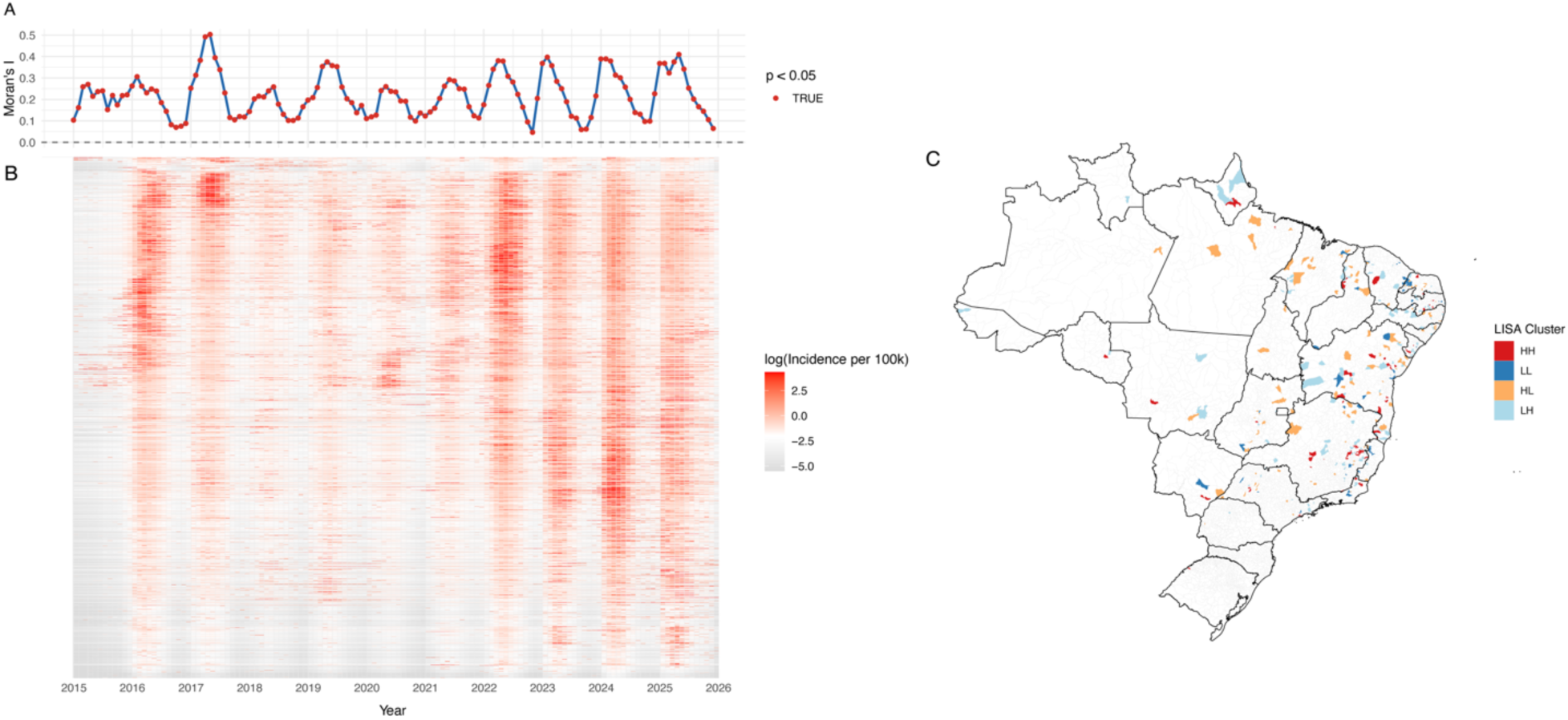
Bayesian spatiotemporal model output and spatial autocorrelation of chikungunya incidence, Brazil, 2015 to 2025. (A) Monthly Global Moran’s I of the space-time interaction term (δ_it_), computed using first-order queen contiguity weights. Values were positive and statistically significant (p < 0·05) in all 132 monthly time steps (range 0·047 to 0·503; median 0·215), indicating persistent structured spatial autocorrelation in municipality-specific deviations beyond the Besag-York-Mollié component. (B) Posterior mean of the structured spatial random effect (u_i_) from the BYM2 parameterization, mapped at the municipality level. Approximately 70% of total spatial variance was attributable to structured geographic clustering (*φ =* 0·70, 95% CrI 0·65 to 0·76). Values ranged from −3·66 to +4·46 on the log scale, corresponding to more than an 80-fold difference in baseline incidence between the most and least affected municipalities. A negative binomial sensitivity analysis yielded highly consistent estimates (*φ* =0·693, 95% CrI 0·641 to 0·744). (C) Municipality-level Local Indicators of Spatial Association (LISA) cluster map derived from local Moran’s I of the space-time interaction term. Municipalities are classified as high-high (HH; red, a municipality with above-average incidence surrounded by above-average neighbors), low-low (LL; dark blue, below-average surrounded by below-average neighbors), high-low (HL; orange, above-average surrounded by below-average neighbors), or low-high (LH; light blue, below-average surrounded by above-average neighbors). Only municipalities with statistically significant local clustering (p < 0·05) are shown; unclassified municipalities are left blank. HH clusters are concentrated in the Northeast and parts of the Central-West, consistent with the macro-regional transmission gradients shown in Figure 2.

Geographic decoupling of transmission intensity and case fatality represents a second structural feature of the epidemic, distinct from and reinforcing the demographic stratification of severity. The geographic distribution of severe outcomes did not mirror the distribution of reported transmission, and the discordance deepened across epidemic periods (Table S5). During the first epidemic wave (2016–17), the Northeast concentrated both the highest transmission intensity and the highest macro-regional case fatality among hospitalized cases (6·36%, 186 of 2,926). By the second wave (2024–25), this alignment had broken: the Central-West, which had become the dominant high-transmission region, had a case fatality of 3·54% (81 of 2,285), while the Southeast, where recent transmission intensity was moderate, recorded the highest case fatality at 4·95% (163 of 3,291). Northeast case fatality fell to 1·38% (22 of 1,592) despite the region’s historically severe burden. This progressive spatial discordance between transmission and hospitalized case fatality is the empirical basis for the dual-axis vaccination allocation framework.

The demographic profile of severe outcomes differed sharply from that of reported infections (Figure 4; Tables S2 and S3). Whereas reported cases were consistently concentrated among working-age adults, hospitalization and death rose steeply with age, increasing markedly from age 65 onwards. In Cox proportional hazards models, patients aged 80 years and older had a more than tenfold higher hazard of death after hospitalization compared with adults aged 20 to 29 years (HR 10·57, 95% CI 7·64 to 14·62). Kaplan-Meier curves declined sharply beyond age 65 across all three clinical intervals, whereas survival among younger adults remained high throughout follow-up (Figure 4A, 4B, 4C).

**Figure 4:**
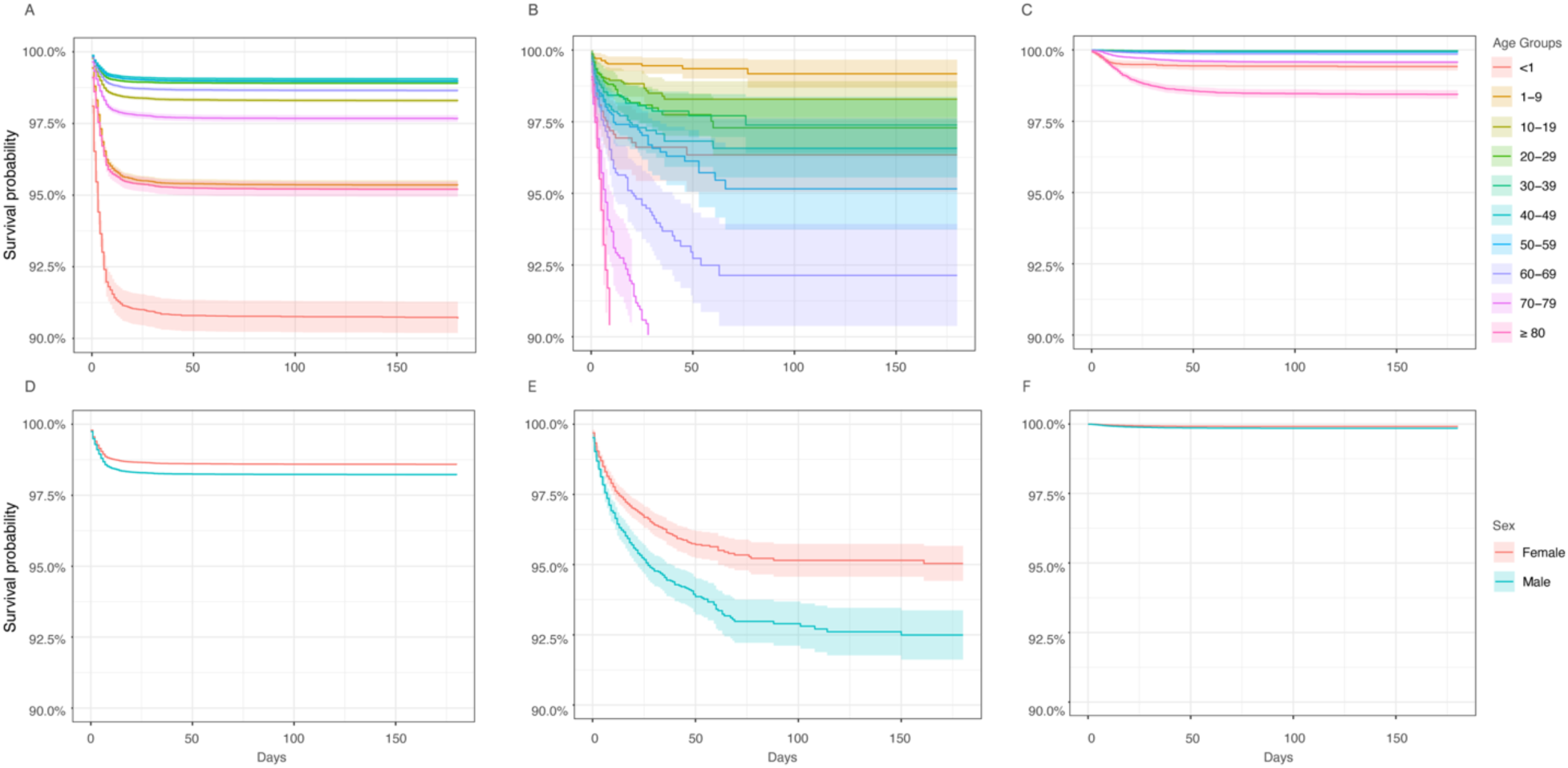
Demographic decoupling of chikungunya transmission and severity, Brazil, 2015 to 2025. Kaplan-Meier survival estimates for three sequential clinical intervals stratified by age group and sex. (A, B, C) Stratification by age group (ten-year intervals; infants as a separate category). Survival declined sharply beyond age 65 across all three intervals. (D, E, F) Stratification by sex. Male progressed faster than female to hospitalization and to death despite contributing fewer reported cases overall. Panels A and D show symptom onset to hospitalization; B and E show hospitalization to death; C and F show symptom onset to death. Shaded bands indicate 95% confidence intervals. Race or ethnicity stratification is shown in Figure S4. Numbers of patients at risk and censored are shown in Table S6.

A parallel divergence by sex was evident. Males progressed faster than females to hospitalization (HR 1·26, 95% CI 1·22 to 1·29), to death after hospitalization (HR 1·47, 95% CI 1·28 to 1·70), and to death from symptom onset (HR 1·76, 95% CI 1·59 to 1·96), despite contributing a smaller share of reported cases, a pattern indicating both sex-differential ascertainment for infection and higher severity risk among males. Race or ethnicity differences were most pronounced in time to hospitalization rather than in post-hospitalization survival. Indigenous individuals showed faster progression to hospitalization (HR for onset to hospitalization 1·49, 95% CI 1·21–1·84), and white individuals showed higher hazards across all three intervals relative to Pardo individuals (HR for onset to hospitalization 1·37, 95% CI 1·33–1·42. HR for onset to death 1·47, 95% CI 1·29 to 1·67; Figure S4). These gradients were concordant in direction and approximate magnitude in mutually adjusted Cox models (Table S3, appendix), indicating they are not artefacts of confounding among age, sex, and race or ethnicity. Clinical presentation was dominated by overlapping symptom combinations rather than isolated features, with fever, myalgia, headache, and arthralgia the most frequently reported symptoms (Figure 5). This symptom-age covariation is consistent with SINAN notifications being a biased sample of infections weighted toward care-seeking working-age adults, which is why notifications cannot substitute for mortality data in identifying who is at highest risk. The divergence between the age profile of reported cases and that of severe outcomes thus reflects both surveillance ascertainment bias and genuine age-dependent differences in disease severity. Municipality-level symptom-onset-to-hospitalization delays were longest in the North and Central-West (Figure S5), consistent with the geographic expansion of transmission into areas with lower diagnostic capacity (Figure S6).

**Figure 5.**
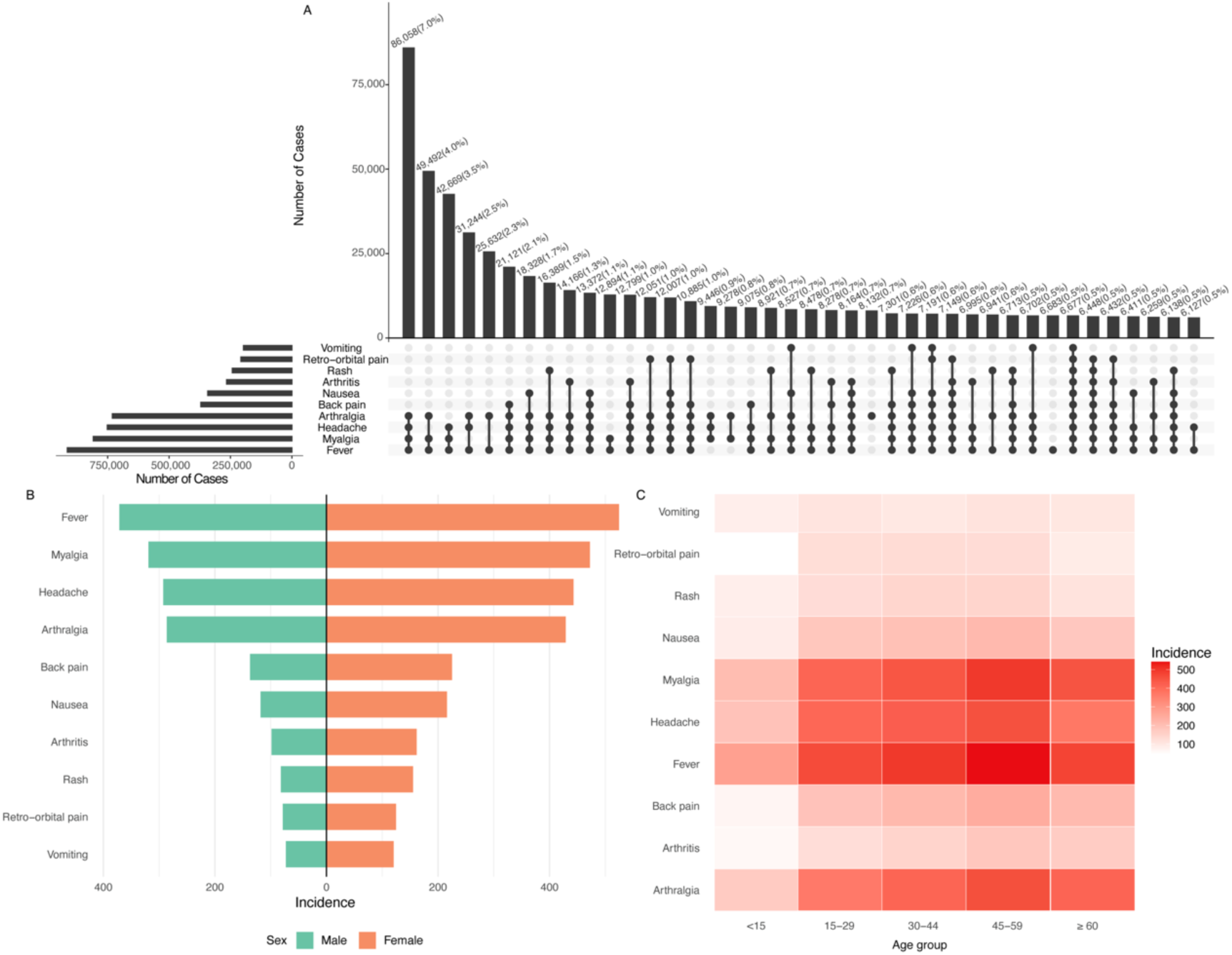
Symptom co-occurrence and demographic distribution of reported clinical manifestations among laboratory-confirmed chikungunya cases, Brazil, 2015 to 2025. (A) UpSet plot showing the frequency of the most common symptom combinations among reported cases. Horizontal bars indicate the total number of cases reporting each individual symptom; vertical bars indicate the size of each unique symptom intersection. Fever, myalgia, headache, and arthralgia were the most frequently reported symptoms, and the largest intersections corresponded to their co-occurrence, indicating that most cases presented with overlapping systemic features rather than isolated manifestations. (B) Population-standardized incidence of individual symptoms by sex. Incidence was higher among females than males for all major symptoms, consistent with higher notification rates among females. Sex categories are shown in the same color palette as figure 1D and figure 4. (C) Population-standardized incidence of individual symptoms by age group. Core systemic symptoms peaked among adults aged 30 to 59 years, mirroring the age profile of reported infections.

A rule that prioritizes municipalities by transmission intensity alone recovers 97% of 2024–25 reported cases but misses most municipalities with high proportions of older adults at elevated mortality risk. An age-only rule captures the elderly population but leaves 82% of recent cases in municipalities it would not prioritize (Figure S9). Neither the spatial gradients in Figures 2 and 3 nor age structure alone therefore captures both dimensions, and a joint targeting rule is required (Figure S9). We translate the two axes into a single municipality-level allocation framework (Figure 6). Each of Brazil’s 3,879 municipalities with model estimates was placed into a joint empirical tertile of recent (2024–25) posterior incidence rate ratio and proportion of population aged 65 years or older, yielding three policy zones: transmission-control priority (predominantly Central-West and parts of the North), clinical-preparedness priority (predominantly Southeast and South), and combined-priority (predominantly Northeast and Southeast). Tertile thresholds were chosen as a pre-specified, data-driven discretization that avoids hand-picked numeric cut-offs while balancing interpretability and operational simplicity. The equal zone counts for the transmission-control and clinical-preparedness groups (each n=832) are a mechanical consequence of this construction, not an empirical finding; the macro-regional asymmetry within each group is the substantive result. The framework is intentionally designed as a planning heuristic, updatable as surveillance data accumulate. The remaining municipalities fell into intermediate (blended) cells of the joint tertile classification. Central-West municipalities were predominantly classified as transmission-control priority (74·3%, 208 of 280), consistent with the 2024–25 epidemic expansion into a region with a younger demographic structure. Southeast and South municipalities were predominantly classified as clinical-preparedness priority (46·9%, 395 of 842 and 77%, 171 of 222, respectively), reflecting older population age structure against lower recent transmission intensity. The Northeast contributed the largest share of combined-priority municipalities (16·3% of its classified municipalities, n=115), consistent with its role as both a long-standing endemic region and a population aging more rapidly than the national average. A time-window sensitivity analysis using cumulative 2015–25 IRRs confirmed that the macro-regional classification was stable to the choice of surveillance period (Figure S7); Figure S9 demonstrates the impossibility of recovering both dimensions from single-axis rules, and Figure S8 provides the corresponding decision-maker reference.

**Figure 6:**
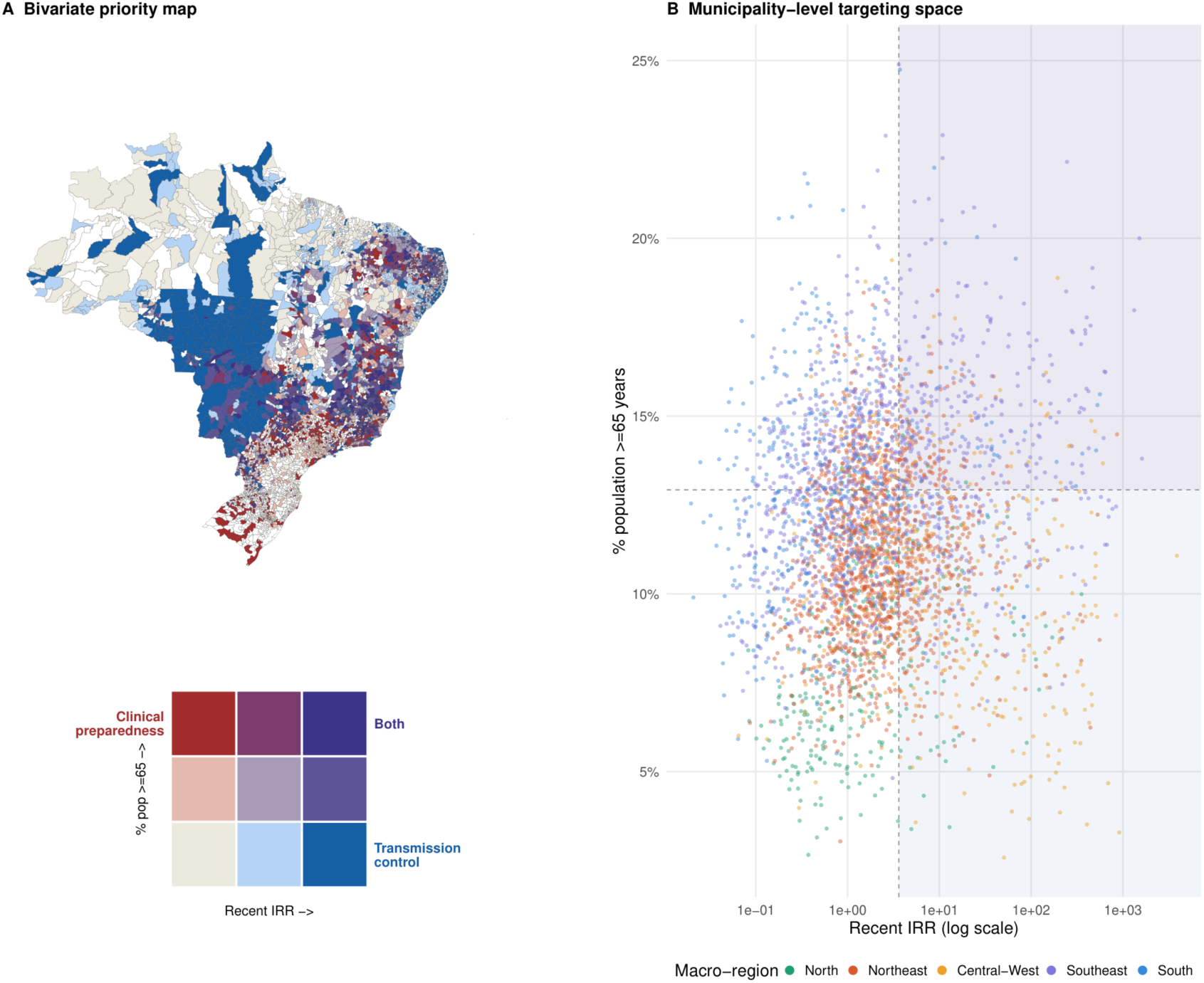
Municipality-level allocation rule for chikungunya vaccination in Brazil, 2024–25. A single rule combining two axes of risk (recent transmission intensity and population age structure) classifies each of Brazil’s 3,879 municipalities with model estimates into one of three policy zones. Tertile thresholds were chosen to balance interpretability and operational simplicity; the framework is a planning heuristic intended to be updated as surveillance data accumulate, and a sensitivity analysis using cumulative 2015–25 IRRs yields a broadly concordant regional classification (Figure S7). **(A) Bivariate priority map.** Color encodes the joint empirical tertile of (i) posterior mean incidence rate ratio (IRR) relative to the national municipality-median in 2024–25, from the Bayesian spatiotemporal model, and (ii) the proportion of the municipality population aged 65 years and older from the IBGE 2022 census. The three corner colors identify distinct policy priorities: blue (high transmission, lower elderly share) indicates transmission-control priority; red (lower transmission, higher elderly share) indicates clinical-preparedness priority; and purple (high on both dimensions) indicates combined-priority. Intermediate cells represent blended zones. Tertile thresholds: IRR 1·03 and 3·61; proportion aged 65 and older 10·4% and 12·9%. **(B) Municipality-level targeting space.** Each point represents one municipality, colored by macro-region. The x-axis (recent IRR, log scale) and y-axis (proportion aged 65 and older) reproduce the two dimensions from panel A; background shading reproduces the three policy zones and dashed lines mark upper tertile thresholds. Municipality counts at displayed thresholds: transmission-control priority, n = 832 (predominantly Central-West and parts of the North); clinical-preparedness priority, n = 832 (predominantly Southeast and South); combined-priority, n = 461 (predominantly Northeast and Southeast). Equal group sizes for the transmission-control and clinical-preparedness zones are a mechanical consequence of the tertile construction; the macro-regional asymmetry within each group, not the group counts, is the substantive finding (Table S7).

## Discussion

Chikungunya in Brazil is spatially structured and demographically stratified, and the two structures are not congruent. Spatial clustering was high and stable throughout the surveillance period, spanning annual outbreaks from 2016 to 2025 with major epidemic peaks in 2016–17 and 2023–24, robust to distributional assumptions and independent of national Zika co-circulation trends. The 2024–25 period produced the largest relative shift in model-estimated transmission intensity toward the Central-West. Demographically, reported infections concentrated among working-age adults, whereas older adults and males bore the highest risks of hospitalization and death. Our findings resolve a gap that prior national analyses left open: neither the state-level characterization of temporal and demographic burden nor the concurrent cohort of hospitalization and death risk addressed whether severity gradients vary geographically or how they translate into a municipality-level allocation rule.^12,14^ The present analysis integrates both dimensions across the full 2015–25 surveillance period and produces the framework the ongoing SUS rollout requires, with direct consequences for IXCHIQ deployment under a safety signal concentrated in the group bearing the highest mortality risk.

The spatial pattern persisted after conditioning on national Zika incidence trends; however, this adjustment does not exclude shared surveillance pressure operating at the municipality level. This extends the most comprehensive prior nationwide analysis, which described recurrent regional outbreaks through 2022 but predated the Central-West expansion and did not test independence from Zika.^11^ It is also consistent with phylogeographic evidence identifying the Northeast as a recurrent source of interregional viral exchange and with ecological work linking chikungunya incidence to social vulnerability and local population structure.^10^ The Northeast-to-Central-West reconfiguration may reflect a combination of spatial heterogeneity in population immunity, differential population susceptibility from lower prior exposure, urbanization patterns, and continued interregional virus exchange, rather than a single dominant mechanism.

The second main finding is the demographic decoupling between notified cases and severe outcomes. Surveillance captures only a small fraction of infections, mainly among care-seeking adults with laboratory confirmation, so the age distribution of notified cases reflects healthcare contact as much as infection. The concentration of notified cases in adults aged 25–55 likely reflects higher symptomatic burden and care-seeking. Infections in children and adolescents are probably underrepresented, as CHIKV infection in younger age groups more frequently presents as mild or asymptomatic disease, reducing health-seeking behavior and the probability of laboratory confirmation^25^. That distinction helps reconcile the concentration of reported cases in adults aged 25 to 55 years with the steep age gradient in mortality observed here and with patterns inferred in national modeling.^12^ The elevated mortality observed among infants is consistent with the U-shaped severity pattern reported elsewhere, while not capturing deaths that may occur before hospitalization.^12^

The sex and race or ethnicity findings point to a mixture of differential ascertainment and differential progression once disease is clinically apparent. The female predominance among notifications with faster male progression to hospitalization and death is consistent with sex differences in care seeking and confirmation combined with higher comorbidity burden in older males, and possibly greater household *Aedes aegypti* exposure among females.^12,13,26^ Male excess is more evident in progression to severe outcomes once disease is clinically apparent. Race or ethnicity gradients in time to hospitalization, notably faster progression among Indigenous individuals and a white–Pardo differential that likely reflects ascertainment differences in higher-resource municipalities rather than biological susceptibility, are reported in Figure S4 and Table S3 and should be interpreted with caution given 23·2% missing records.^14,22,26,27^

These spatial and demographic structures have direct implications for the ongoing chikungunya vaccine rollout through the SUS and the *Programa Nacional de Imunizações*. The persistent clustering and the relative shift in transmission intensity toward the Central-West during 2024–25, against a background of ongoing outbreaks across all Brazilian macro-regions, suggest that residual susceptibility is concentrated in areas with lower cumulative incidence and rising transmission, particularly the Central-West and parts of the North. The macro-regional case fatality data reinforce this distinction: despite leading transmission intensity in 2024–25, the Central-West recorded lower case fatality among hospitalized cases (3·54%, 81 deaths / 2,285 hospitalized) than the Southeast (4·95%, 163 deaths / 3,291 hospitalized), the reverse of the 2016–17 alignment when the Northeast simultaneously led both transmission and the highest chikungunya mortality burden, with Ceará alone accounting for 30·6% of all suspected deaths nationally between 2015 and 2023. ^28^ This reversal is mechanistically coherent with the individual-level survival findings, the 2024–25 Central-West epidemic expanded primarily into working-age adults, the demographic sustaining the highest reported case burden, whereas the Southeast’s older population age structure directly maps onto the tenfold age gradient in post-hospitalization mortality documented in the Cox models. The geographic decoupling of CFR from incidence is therefore not a paradox but a predictable consequence of demographic structure, and it is precisely this spatial-demographic incongruence that makes a uniform age-targeted vaccine rule inadequate. Figures 6 and S7 translate those mapped gradients into the allocation framework, and Figure S8 into action. Because the populations driving reported transmission are not the same as those at highest risk of death, a strategy that combines geographic prioritization with age-stratified clinical preparedness follows directly from these findings.^12,14^ An age-targeted rule alone would leave 82% of 2024–25 reported cases in municipalities it would not prioritize; the transmission axis largely closes that gap in surveillance data without displacing the clinical-preparedness logic for older adults.

The municipality-level allocation framework translates that decoupling into an implementable policy rule. Transmission-control priority municipalities, concentrated in the Central-West and parts of the North, are the places where preemptive vaccination of younger and middle-aged adults could help blunt epidemic expansion (Figure S8). This age profile aligns directly with the current ANVISA label for IXCHIQ, making transmission-control zones the most operationally actionable for immediate preemptive rollout. Clinical-preparedness priority municipalities, concentrated in the Southeast and South, are where lower recent transmission coincides with older age structure, making hospital readiness especially important. Combined-priority municipalities, concentrated in the Northeast and Southeast, require both functions. A sensitivity analysis using cumulative IRRs yielded a broadly concordant classification (Figure S7), and the three zones in Figure 6 operationalize the patterns mapped in Figures 2 and 3.

The post-licensure IXCHIQ safety signal in adults aged 60 years and older constrains, but does not uniformly resolve, the targeting question for older adults in high-endemic settings. The US FDA suspended IXCHIQ’s biologics license in August 2025 following serious adverse events, including one encephalitis death and over 20 cases of vaccine-strain chikungunya-like illness, predominantly in adults aged 60 years and older with underlying chronic conditions.^1,2,4,29^ Regulatory authorities in the EU (EMA), Brazil (ANVISA), and Canada did not revoke licensing but instead revised prescribing guidance to require individual benefit–risk assessment before vaccinating older adults, recognizing that high endemic background risk may shift the balance toward selective use. The FDA safety signal arose in travelers vaccinated in low-transmission settings, where the risk of chikungunya is low and vaccine-associated harm is difficult to justify; Brazil’s endemic context, with substantially higher exposure risk, argues against blanket exclusion in favor of restricted use in municipalities where high transmission and older age structure coincide. This refines the targeting problem identified in prior vaccine impact and cost-effectiveness analyses: the older adults at highest mortality risk concentrate in the Southeast and South, where recent transmission is lower, while the rising-incidence Central-West front is demographically young. Combined-priority municipalities, where high transmission and older age structure coincide, highlight the most consequential gap: the lack of a CHIKV vaccine safe for adults aged 60 and older, underscoring the need to understand adverse-event mechanisms in this group and to develop suitable vaccine platforms. Forthcoming evidence on Vimkunya and Butantan-Chik may further reshape this balance if their platforms differ in older adults.^3^

The findings also reveal geographic heterogeneity in diagnostic and clinical capacity that introduces inequities in how chikungunya burden is measured, who receives timely care, and ultimately who benefits from any control intervention. Municipality-level heterogeneity in laboratory confirmation and in delays from symptom onset to hospitalization, particularly in the North and Central-West, signals persistent inequities in access to diagnosis and clinical care, reflecting both capacity constraints and demand-side barriers such as health-seeking behavior and transportation. The female predominance among notified cases, race or ethnicity differences in time to hospitalization, and the under-ascertainment of race or ethnicity in lower-confirmation municipalities together indicate that vaccination alone will not resolve disparities in access, the same barriers that delay diagnosis and hospitalization will also impede vaccine uptake, risking coverage that replicates rather than corrects existing inequities. Expanding point-of-care diagnostic coverage in lower-confirmation municipalities, leveraging the existing integration of chikungunya within dengue surveillance, and conducting representative serosurveys would reduce surveillance ascertainment bias, improve targeting, and support more equitable vaccine deployment.^14,30^

These findings provide the first municipality-level evidence base for the ongoing SUS rollout, but several limitations bear on interpretation. Surveillance captures only a fraction of infections, and spatial heterogeneity in confirmation rates and potential disruptions during 2020–2021 may partly confound regional gradients. These features introduce the potential for selection and information bias in both incidence and severity estimates, which we address by focusing on relative gradients, conducting sensitivity analyses, and interpreting the allocation framework as a heuristic rather than a direct measure of true incidence.^30^ Macro-regional case fatality estimates (Table S5) are additionally sensitive to regional differences in hospitalization thresholds and death ascertainment and should be interpreted as reflecting the joint distribution of disease severity and health-system capacity rather than biological lethality. Because notified cases reflect care seeking and diagnostic access, the demographic profile of notified disease is not equivalent to the infected population, and inference about who sustains transmission remains indirect. Clinical diagnostic overlap with dengue can also produce misclassification during co-circulation, especially in the acute phase when symptom profiles converge,^29^ supporting fuller utilization of the existing integrated chikungunya-dengue surveillance infrastructure. National monthly Zika incidence may not fully capture municipality-level arboviral co-circulation, and late 2025 SINAN records may still be subject to updating. Mobility, vector indices, longitudinal linkage, and representative serology were not available and would improve future predictive work.

With IXCHIQ already deployed through the SUS and the epidemic front continuing to shift, the municipality-level framework presented here offers an immediately actionable, updatable basis for targeting, one that neither a geography-blind nor an age-only rule can provide.

## Contributors

QZ curated the data, conducted the statistical analyses, and prepared the figures. SC and AIB conceived and designed the study, supervised the analyses, and wrote the first draft manuscript. QZ, SC and AIB had full access to all the data and contributed to interpretation of the findings. FSB, FA, WMS provided expert contextualization of arboviral dynamics in Brazil, commented and provided edits to the manuscript. All authors approved the final version.

## Data sharing

This study used anonymized, publicly available surveillance data from SINAN (DATASUS) and IBGE. All processed data and analysis code are available at https://github.com/hemaanizg/Chikungunya-Spatiotemporal-Analysis. Original SINAN microdata are publicly available at https://datasus.saude.gov.br/transferencia-de-arquivos/.

## Acknowledgments

We acknowledge the Brazilian Ministry of Health and the DATASUS team for maintaining and curating SINAN, and the *Instituto Brasileiro de Geografia e Estatística* (IBGE) for providing population and boundary data. We thank Dr. Livia Vinhal Frutuoso, Dr Sulamita Barbiratto and Dr. Marcela Santos from the Arboviral Surveillance unit in the Brazilian MoH for their comments and insights. We thank colleagues in the Bento lab for helpful comments on the manuscript, and colleagues at the Cornell Atkinson Center for Sustainability for discussion and support. We are deeply grateful for the extraordinary public stewardship of these national data systems, which made this work possible. The views expressed are those of the authors and do not necessarily reflect those of the Brazilian Ministry of Health and IBGE.

## Supplementary Materials

### Supplementary Methods

#### Data sources and study population

We performed a nationwide retrospective analysis of laboratory-confirmed chikungunya virus (CHIKV) cases reported in Brazil between January 2015 and December 2025. Individual-level surveillance records were obtained from the *Sistema de Informação de Agravos de Notificação* (SINAN) via DATASUS, the information technology department of Brazil’s Unified Health System, the *Sistema Único de Saúde.*^1^ SINAN is the mandatory notification system for infectious diseases in Brazil and includes patient demographics such as age, sex, race or ethnicity, municipality of residence, dates of symptom onset, notification, and hospitalization, reported clinical manifestations, and final outcomes.

Analyses were restricted to laboratory-confirmed chikungunya cases as defined by SINAN case classification criteria, and suspected or discarded notifications were excluded. Population denominators stratified by age and sex were obtained from intercensal estimates provided by the Brazilian Institute of Geography and Statistics.^2^ Municipality boundary files from IBGE were used to define spatial adjacency for subsequent spatial analyses.^21^This restriction improves specificity but substantially reduces case counts relative to the total burden, because laboratory confirmation rates for chikungunya in Brazil have been estimated at 13% to 62% of suspected cases. Consequently, these analyses characterize the epidemiology of laboratory-confirmed disease^1,3^ and may underestimate true transmission intensity, particularly in municipalities with limited diagnostic capacity. Laboratory confirmation rates for chikungunya in Brazil have been estimated at 13% to 62% of suspected cases,^1,4^ so these analyses characterize the epidemiology of laboratory-confirmed disease and may underestimate true transmission intensity, particularly in municipalities with limited diagnostic capacity.

#### Data preprocessing and variable definition

Records with missing or invalid information on age or sex were excluded. Age was analyzed both continuously and in grouped categories. For survival analyses, age was grouped in ten-year intervals, with infants younger than 1 year as a separate category. For symptom analyses, broader age categories were used: younger than 15, 15 to 29, 30 to 44, 45 to 59, and 60 years or older.

Race or color was recorded in SINAN according to the IBGE self-declaration standard, which classifies individuals into five categories: *branca* (White), *preta* (Black), *amarela* (East Asian), *parda* (mixed ancestry, predominantly European-African-Indigenous combinations), and *indígena* (Indigenous).^5,6,7^ The term pardo refers to individuals of mixed ancestry, predominantly European-African-Indigenous combinations, as defined by the IBGE self-declaration standard.^7^ Following standard practice in Brazilian epidemiological research, preta and parda were aggregated into a single Black category for some analyses of racial disparities.^7^ Race or color was missing or recorded as ignored in 23.2% of confirmed case records; these individuals were excluded from race-stratified analyses but retained in all other analyses. Because missingness was more common in municipalities with lower laboratory-confirmation rates, race-specific estimates may understate inequities in access to care.

Municipality-level incidence, hospitalization, and death rates were calculated as the number of events per 100,000 population, aggregated at the temporal resolution appropriate to each analysis. For analyses of disease progression, three time-to-event intervals were defined in days: symptom onset to hospitalization, hospitalization to death, and symptom onset to death. Individuals who did not experience the event of interest during follow-up were right censored at the last observed date.

#### Descriptive epidemiological analyses

National and regional temporal trends were summarized using weekly aggregated case counts and population-standardized incidence rates. Seasonal patterns were explored using weekly moving averages and deviations from long-term weekly means. Annual age and sex distributions of reported cases were summarized using boxplots. Differences in incidence across race and ethnicity groups were assessed using annual incidence rates and proportional distributions. Municipality-level sex differences in case burden were evaluated by comparing the proportion of female and male cases, with statistical significance assessed using binomial tests and visualized using volcano-style plots.

#### Wavelet analysis of temporal periodicity

To characterize the temporal periodicity of chikungunya transmission, we applied a continuous wavelet transform to national monthly chikungunya case counts from January 2015 to December 2025. Monthly case counts were aggregated at the national level based on notification dates. Wavelet power spectra were computed using the Morlet wavelet function, which provides balanced resolution in time and frequency and is widely used for epidemiological time-series analysis.^23^ Periods ranging from 2 months to 5 years were evaluated. Statistical significance of wavelet power was assessed against an AR(1) red-noise background spectrum, following Torrence and Compo (1998).^8^ The AR(1) model represents the null hypothesis that observed power at each period reflects autocorrelated background noise rather than a genuine periodic signal. Significance contours were drawn at the 1% level, indicating periods where wavelet power significantly exceeded the red-noise expectation. To account for boundary effects inherent to finite time series, results outside the cone of influence were excluded from interpretation. Wavelet analyses were performed in R using the *WaveletComp* package.^6^

#### Clinical symptom co-occurrence

To describe the clinical heterogeneity of reported cases, we examined the co-occurrence of reported symptoms among chikungunya cases. Common symptoms included fever, arthralgia, myalgia, headache, rash, vomiting, conjunctivitis, and leukopenia. Overlapping symptom profiles were visualized using an UpSet plot framework to highlight common multi-symptom combinations.

#### Survival analyses

Time-to-event outcomes were analyzed using Kaplan-Meier methods, stratified by age group, sex, and race or ethnicity, with differences assessed using log-rank tests. Three separate univariable Cox proportional hazards models were fitted, one each for age group, sex, and race or ethnicity, to estimate unadjusted associations between each demographic characteristic and disease progression. Because the aim was to characterize population-level demographic gradients in disease progression rather than to identify independent predictors, Cox models were fitted univariably. Results are reported as hazard ratios with 95% confidence intervals. Proportional hazards assumptions were evaluated using Schoenfeld residuals.

#### Municipality-level delay analyses

Spatial heterogeneity in disease progression and surveillance was assessed by calculating municipality-level median delays for key intervals, including symptom onset to notification and symptom onset to hospitalization. Median values were used to reduce sensitivity to extreme observations. Municipalities with insufficient numbers of events were excluded. Results were visualized using choropleth maps.

#### Bayesian spatiotemporal model

Municipality-level chikungunya case counts were analyzed using a Bayesian hierarchical spatiotemporal model implemented within the Integrated Nested Laplace Approximation framework. Let *Y*_*it*_ denote the number of reported chikungunya cases in municipality during month. The outcome was assumed to follow a Poisson distribution. A log link function was used, with municipality population size included as an offset.

The linear predictor included an intercept, a standardized national monthly Zika incidence covariate, structured and unstructured spatial effects, a temporal random effect, and a space-time interaction term. The structured spatial component was modeled using the Besag-York-Mollié 2 parameterization,^9^ with an intrinsic conditional autoregressive prior defined on a first-order queen contiguity adjacency structure. The parameter *ϕ* represents the proportion of total spatial variance attributable to the structured spatial component, with values close to 1 indicating strong spatial autocorrelation and values near 0 indicating predominantly unstructured spatial heterogeneity. Temporal variation was captured using a second-order random walk, and residual municipality-specific deviations were modeled through an unstructured space-time interaction term.

To assess whether the space-time interaction term exhibited spatial clustering beyond the structured spatial component, we computed Global Moran’s I for each monthly time step using first-order queen contiguity weights.^8,10^ To characterize persistent clustering at the municipality level, we also computed Local Indicators of Spatial Association using the time-averaged posterior mean of the interaction term. Statistical significance was assessed at *p <0.05*. To capture national-level co-circulation of arboviruses, we incorporated a time-varying covariate representing standardized national monthly Zika incidence. As a sensitivity analysis, the model was refit using a negative binomial likelihood to account for overdispersion in municipality-month case counts.^25,26^ Full model specification:

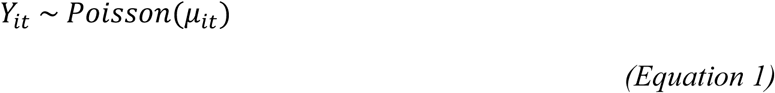

A log link function was used, with municipality population size included as an offset (Equation 2):

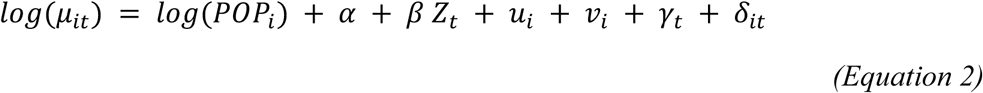

The linear predictor represents the log incidence rate. Here, *μ*_*it*_ denotes the expected number of cases and *POP*_*i*_ represents the municipality population size included as an offset. The parameter *α* represents the overall intercept. The coefficient *β* represents the association between chikungunya incidence and the standardized national monthly Zika incidence covariate *Z*_*t*_. Under this log-link model, exp(*β*) represents the multiplicative change in the chikungunya incidence rate associated with a one-standard-deviation increase in national Zika incidence, conditional on spatial, temporal, and spatiotemporal random effects. As a sensitivity analysis, we refit the model using a negative binomial likelihood to account for overdispersion (Table S4).

#### Spatial effects

Spatial variation was modeled using the Besag-York-Mollié 2 (BYM2) parameterization.^9^ The spatial random effect was decomposed into a structured component *u*_*i*_and an unstructured component *v*_*i*_. The structured spatial component *u*_*i*_ was modeled using an intrinsic conditional autoregressive (ICAR) prior defined on a first-order queen contiguity adjacency structure, such that municipalities sharing a common boundary or vertex were considered neighbors. The parameter *φ* represents the proportion of total spatial variance attributable to the structured spatial component, with values close to 1 indicating strong spatial autocorrelation and values near 0 indicating predominantly unstructured spatial heterogeneity. This specification induces spatial smoothing by allowing geographically adjacent municipalities to exhibit similar underlying risk levels. The unstructured spatial component *v*_*i*_ captures spatially independent heterogeneity.

#### Temporal and space-time effects

Temporal variation was captured by *γ*_*t*_, modeled using a second-order random walk (RW2) to impose smoothness in the temporal trend. Residual variability not explained by the additive spatial and temporal components was modeled using an unstructured space-time interaction term *δ*_*it*_, allowing municipality-specific deviations from the overall temporal pattern.

#### Spatial autocorrelation analyses

To assess whether the space-time interaction term *δ*_*it*_ exhibited spatial clustering beyond the structured BYM2 component, we computed the Global Moran’s I statistic for each monthly time step. For month *t*, let *x = {δ*_*it*_*}* denote the vector of posterior means across all municipalities. The Global Moran’s I is defined as (Equation 3):

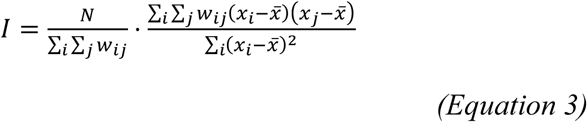

Where *N* is the number of municipalities, *w*_*ij*_ are spatial weights from the first-order queen contiguity adjacency matrix (where *w*_*ij*_ = 1 if municipalities *i* and *j* share a common boundary or vertex, and 0 otherwise), *i* and *j* index individual municipalities and *x̄* is the spatial mean of *δ*_*it*_ at time *t*. Statistical significance was assessed under the assumption of normality, with *p* < 0.05 as the threshold.

To characterize persistent spatial clustering at the municipality level across the full study period, we additionally computed Local Indicators of Spatial Association (LISA) using the Local Moran’s I statistic (Equation 4):

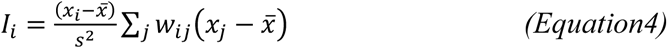

Where *s²* is the variance of *x*. LISA was applied to the time-averaged posterior mean of *δit*. Municipalities were classified into cluster types (HH, LL, HL, LH) based on the sign of *I*_*i*_ and the spatial lag *Σ w*_*ij*_ (*x*_#_ − *x̄*) with significance defined at p < 0.05. All spatial autocorrelation analyses were performed using the spdep package in R.

#### National Zika covariate

To capture national-level co-circulation of arboviruses, we incorporated an external time-varying covariate *Z*_*t*_representing standardized national monthly Zika incidence. Let *C_t_^Z^* denote the total number of confirmed Zika cases nationwide in month *t*, and let *P^Z^* denote the total national population obtained by summing municipality-level population estimates. The raw national monthly Zika incidence rate (per 100,000) was computed as (Equation 5):

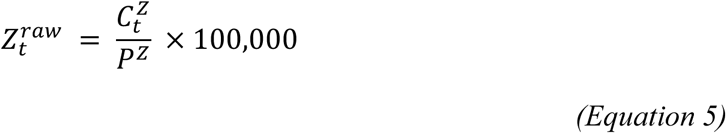

To improve interpretability and numerical stability, the incidence rate was standardized across all months using a z-score transformation (Equation 6):

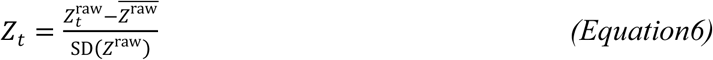

We used national Zika incidence rather than dengue as the arboviral covariate because Zika’s temporally distinct emergence and epidemic peak provided a cleaner test of whether chikungunya spatial gradients simply mirrored broader arboviral surveillance pressure. Dengue’s long-standing endemicity and overlapping seasonality with chikungunya would have introduced strong collinearity, making the covariate difficult to interpret mechanistically. The primary analytic goal was to assess independence of chikungunya spatial structure from co-circulating arboviral dynamics, for which Zika is the more appropriate probe.

#### Incidence rate definition

Rearranging Equation 2, the municipality-level incidence rate can be expressed as (Equation 7):

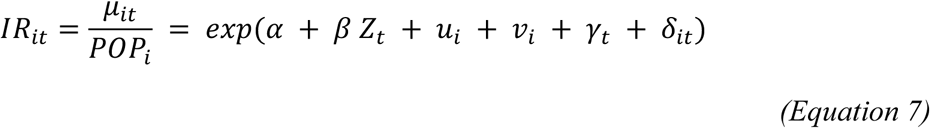

Incidence rates per 100,000 population were obtained by multiplying *IR_it_* by 100,000.

#### Estimated incidence rate ratio definition

To quantify relative transmission intensity across municipalities, we defined a municipality-level estimated incidence rate ratio (*IRR*) as the ratio between the model-estimated municipality incidence rate and the estimated national median municipality incidence rate at the same time point (Equation 8):

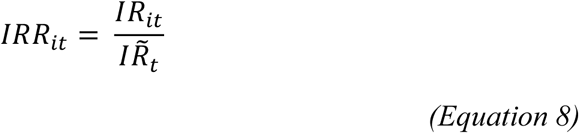

where *IR*_*it*_ denotes the model-estimated incidence rate for municipality *i* at time *t*, and *IR*^H^_*t*_ represents the national median of municipality-level incidence rates at time *t*. Equivalently (Equation 9):

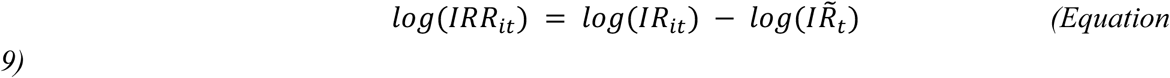

For visualization of regional trajectories, macro-regional summaries were computed as the median *log*(*IRR*) across all municipalities within each macro-region at each time point.

#### Priors and inference

Penalized complexity (PC) priors were specified for precision parameters. Posterior inference was performed using INLA, which approximates marginal posterior distributions via Laplace approximation and numerical integration. Posterior summaries of municipality-level incidence rates were obtained by exponentiating the linear predictor excluding the offset term. All analyses were conducted in R (version ≥4.2). Spatial analyses used the sf and spdep packages, Bayesian spatiotemporal models were fitted using INLA, and survival analyses used survival and survminer. All analyses were version controlled.

#### Municipality-level allocation framework

Each municipality with model estimates was assigned to a joint empirical tertile of recent (2024–25) posterior incidence rate ratio (IRR) and proportion of population aged 65 years or older from the IBGE 2022 census, yielding three policy zones: transmission-control priority (high IRR, lower elderly share), clinical-preparedness priority (lower IRR, high elderly share), and combined-priority (high on both dimensions). As a sensitivity analysis, the framework was reconstructed using cumulative 2015–25 posterior IRRs rather than the 2024–25 window; results are shown in Figure S7.

### Data availability

This study used anonymized, publicly available surveillance data, and ethical approval was not required under Brazilian regulations. All data and code used in this study are available at https://github.com/hemaanizg/Chikungunya-Spatiotemporal-Analysis.

## Supplementary Results

**Figure S1.**
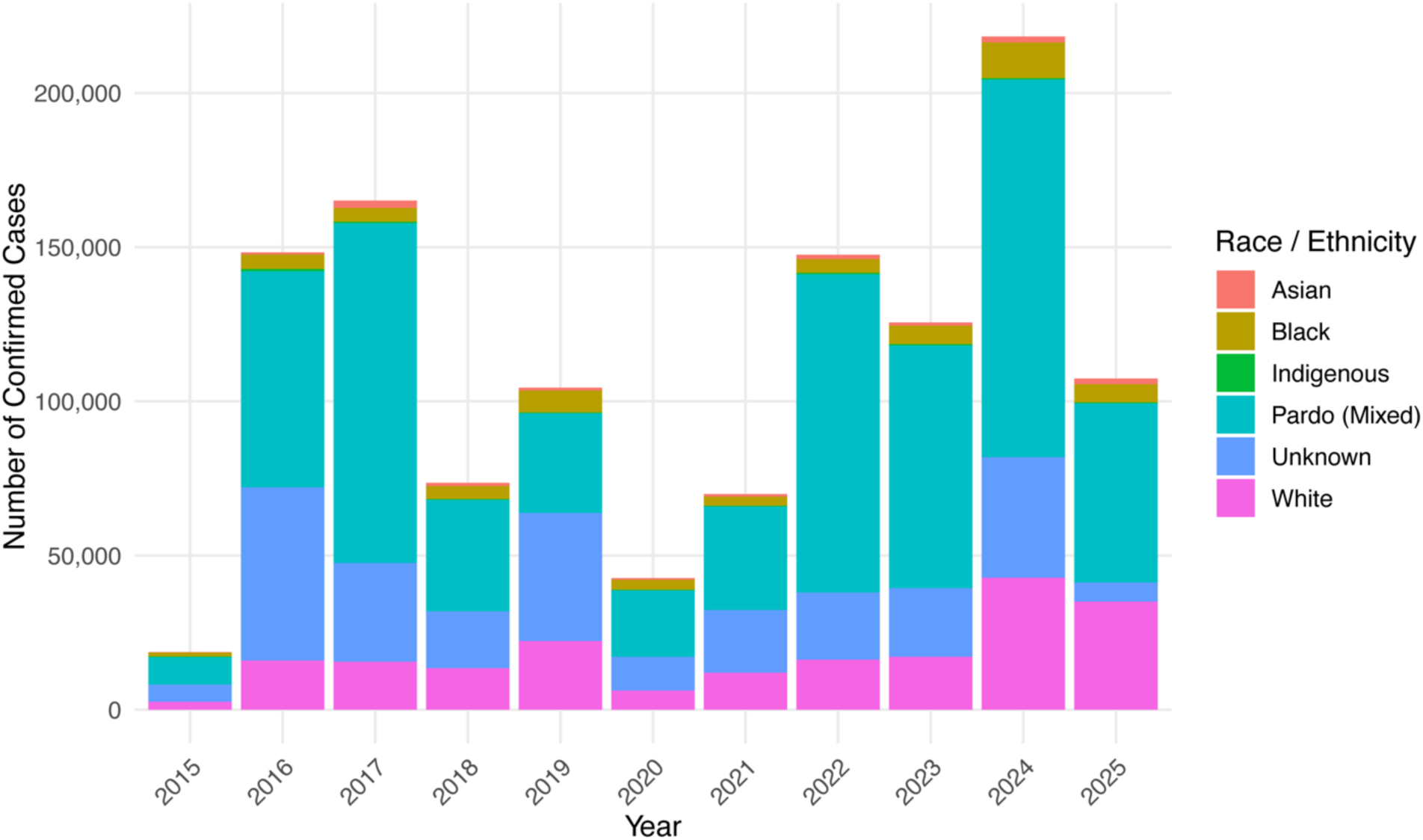
Annual number of confirmed chikungunya cases stratified by race or ethnicity from 2015 to 2025. Individuals classified as Pardo (mixed) consistently accounted for the largest share of reported cases across all years, followed by white and Black populations. Temporal fluctuations in total case counts reflect epidemic waves, while the relative contribution of each ethnic group remained broadly similar over time.

**Figure S2.**
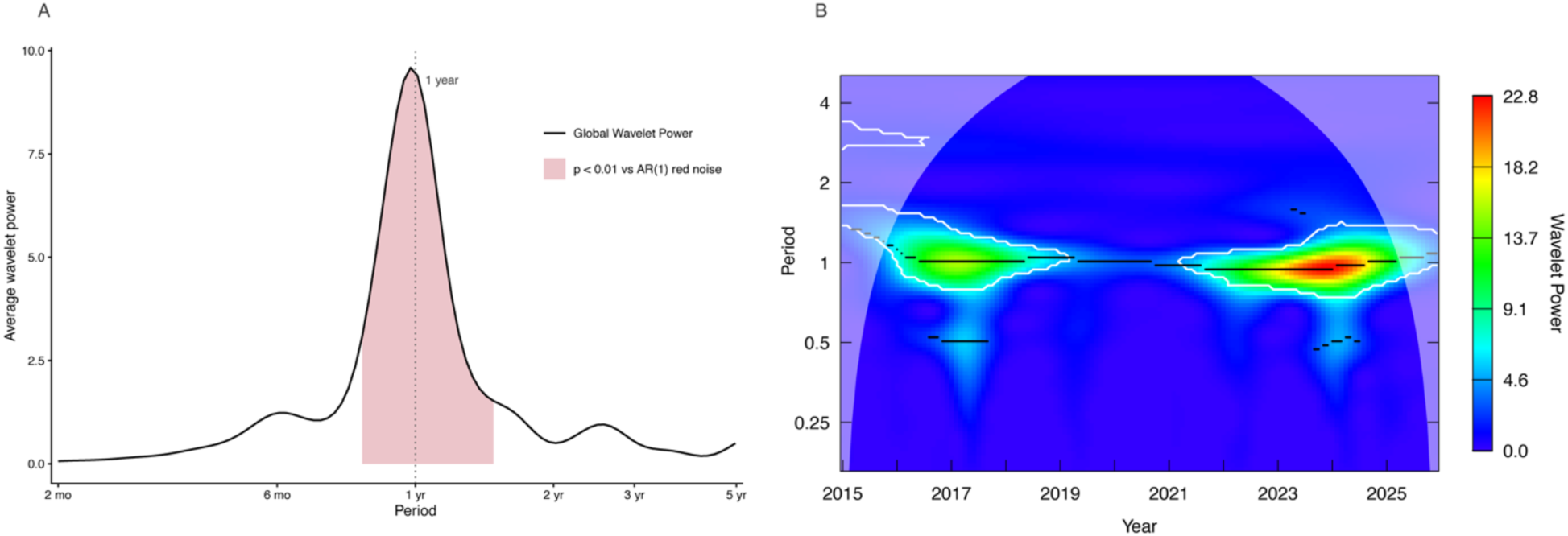
Wavelet power spectrum of national monthly chikungunya case counts, Brazil, 2015 to 2025. The continuous wavelet transform was computed using the Morlet wavelet applied to national monthly aggregated case counts. Color intensity indicates wavelet power at each combination of time (x-axis) and period (y-axis). The black contour encloses regions where power significantly exceeds a red-noise (autoregressive order-one) background at the 1% level, indicating genuine periodic signals rather than autocorrelated noise. A dominant annual cycle is present throughout the study period and is markedly amplified during the 2016 to 2017 and 2023 to 2024 epidemic waves. No sustained multi-year periodicity is evident. The lighter shaded region at the edges indicates the cone of influence, where edge effects limit interpretation.

**Figure S3.**
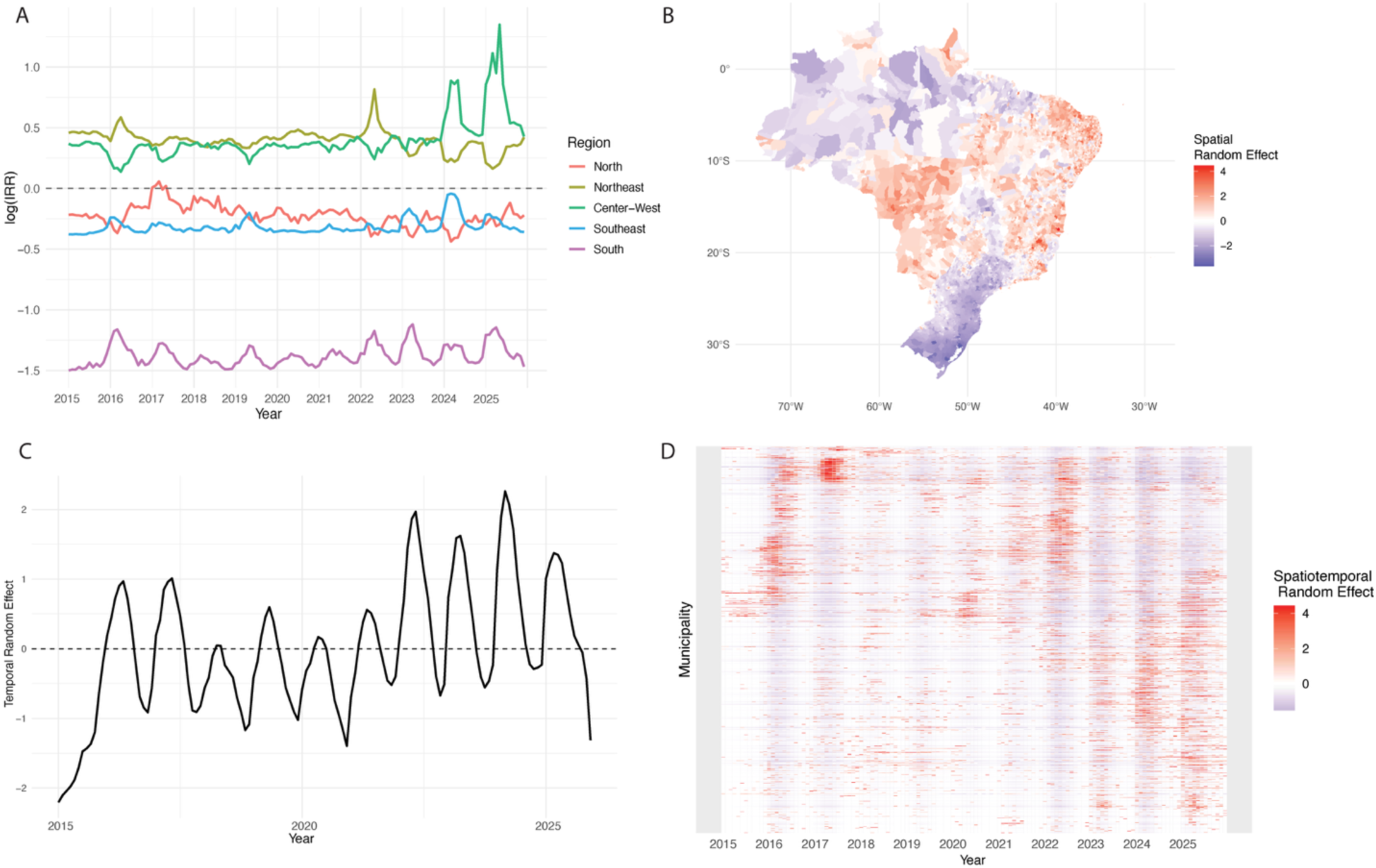
Decomposition of the Bayesian spatiotemporal model. (A) Monthly macro-regional log incidence rate ratios relative to the national municipality-median incidence. (B) Posterior mean of the spatial random effect (BYM2), representing persistent spatial structure after accounting for covariates and temporal trends. (C) Posterior mean of the temporal random effect (RW2), representing the national temporal trend after adjusting for spatial structure. (D) Posterior mean of the unstructured space-time interaction term, visualized as a municipality-by-month heatmap, capturing residual local deviations beyond additive spatial and temporal effects.

**Figure S4.**
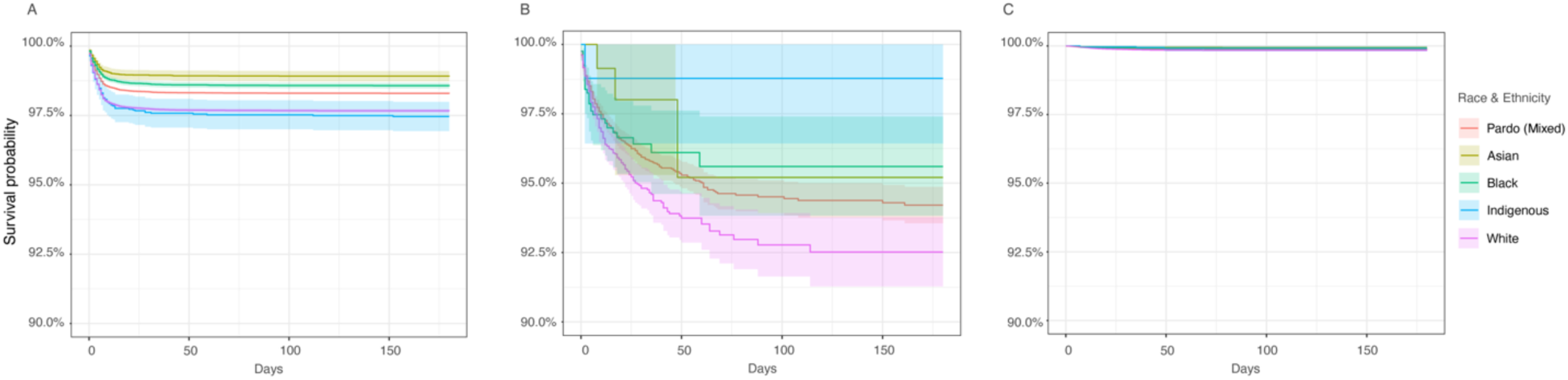
Kaplan-Meier survival estimates by race or ethnicity along the chikungunya clinical pathway, Brazil, 2015 to 2025. Survival estimates for three sequential clinical intervals stratified by race or ethnicity as classified per IBGE self-declaration categories (white, Black, Pardo, Asian, Indigenous). (A) Symptom onset to hospitalization. (B) Hospitalization to death. (C) Symptom onset to death. Individuals with missing race or ethnicity (23.2% of records) were excluded. Differences were most pronounced for time from symptom onset to hospitalization, consistent with inequities in care access rather than biological susceptibility. Shaded bands indicate 95% confidence intervals. Numbers at risk are shown below each panel.

**Figure S5.**
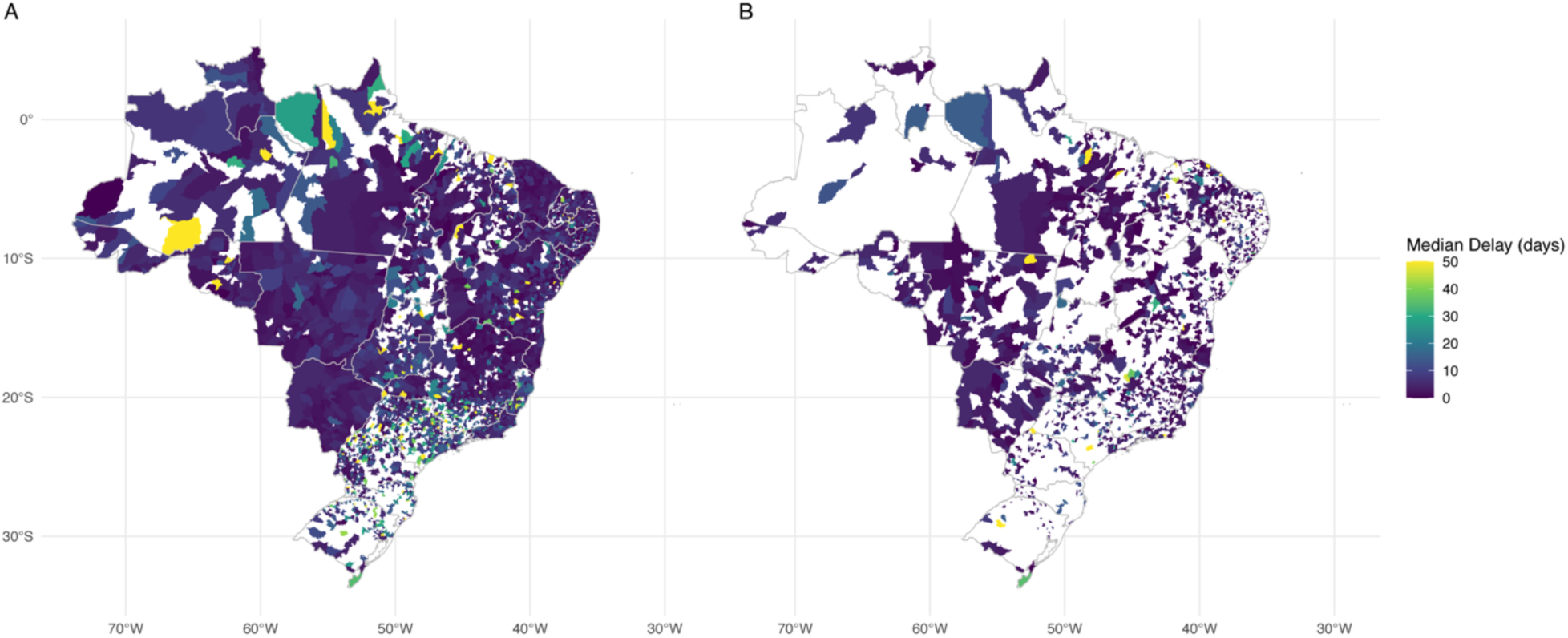
Municipality-level median delays in chikungunya notification and hospitalization, Brazil, 2015 to 2025. Choropleth maps show municipality-level median delays in days for two care-pathway intervals. (A) Symptom onset to notification. Median delays were generally short across most of Brazil but showed clusters of longer reporting times in parts of the North and South, consistent with reduced laboratory-confirmation capacity in those areas. (B) Symptom onset to hospitalization. Geographic heterogeneity was more pronounced, with longer median delays in parts of the North and Central-West. These patterns are consistent with persistent inequities in primary care access and hospital referral pathways across SUS regions. Municipalities with fewer than five events were excluded to reduce instability in median estimates. Values are shown in days.

**Figure S6.**
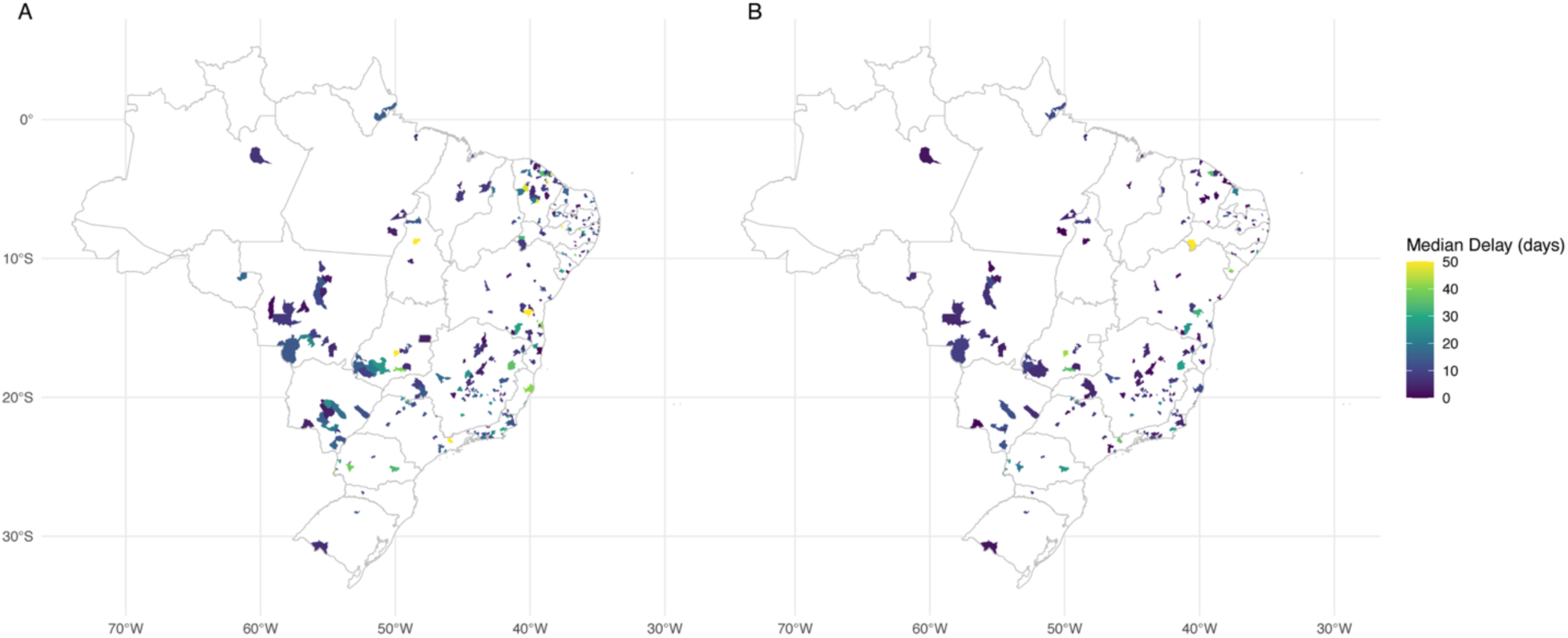
Municipality-level median time to death across the chikungunya clinical pathway. (A) Median time from symptom onset to death could be estimated only in a limited number of municipalities, reflecting the low frequency and uneven geographic distribution of fatal cases. (B) A similar pattern was observed for the median time from hospitalization to death, which was estimable in even fewer municipalities, consistent with the strong attrition of cases along the disease severity pathway.

### Spatial model diagnostics and sensitivity analyses

The primary Poisson spatiotemporal model showed strong convergence and consistent parameter estimates across diagnostic checks. The negative binomial sensitivity analysis yielded a substantially improved model fit by both DIC (484,146 vs 806,237) and WAIC (470,086 vs 1,945,718), confirming overdispersion in municipality-month case counts, but produced highly consistent estimates of the structured spatial fraction (φ = 0.693, 95% CrI 0.641 to 0.744 vs φ = 0.699, 95% CrI 0.649 to 0.755 in the primary model; Table S4). The Zika covariate association was similarly robust across both specifications (exp(β) 1.174, 95% CrI 0.986 to 1.398 in the negative binomial model vs 1.120, 95% CrI 0.943 to 1.331 in the Poisson model). Space-time interaction precision differed between models (0.544 vs 0.143), consistent with the negative binomial capturing overdispersion that the Poisson model partially absorbed into the interaction term; this did not affect the primary conclusions regarding spatial structure or the Zika independence finding. The cumulative IRR-based sensitivity analysis of the allocation framework (Figure S7) produced a macro-regional classification broadly concordant with the 2024–25 window used in the main analysis, supporting the robustness of the targeting framework to the choice of time window.

**Figure S7.**
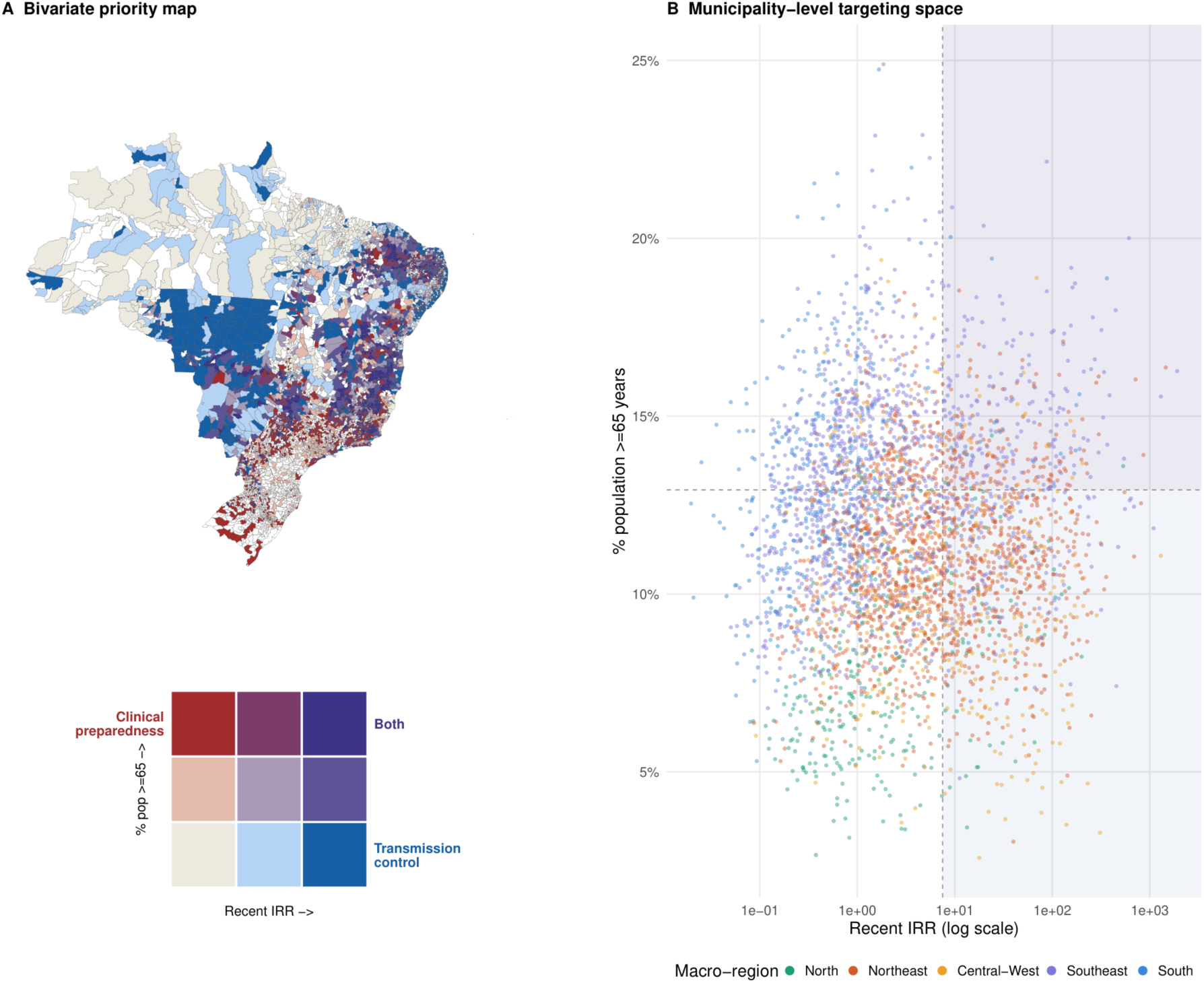
Combined geographic and demographic targeting using cumulative 2015 to 2025 IRRs, Brazil (sensitivity analysis). Sensitivity version of figure 6 constructed using cumulative posterior incidence rate ratios (IRRs) over the full 2015 to 2025 study period rather than the 2024 to 2025 window used in the main analysis. (A) Bivariate priority map and (B) municipality-level targeting space are constructed using identical joint empirical-tertile classification and the same IBGE 2022 proportion of population aged 65 years and older. The macro-regional distribution of priority zones is broadly concordant with figure 6, supporting the robustness of the targeting framework to the choice of time window.

**Figure S8.**
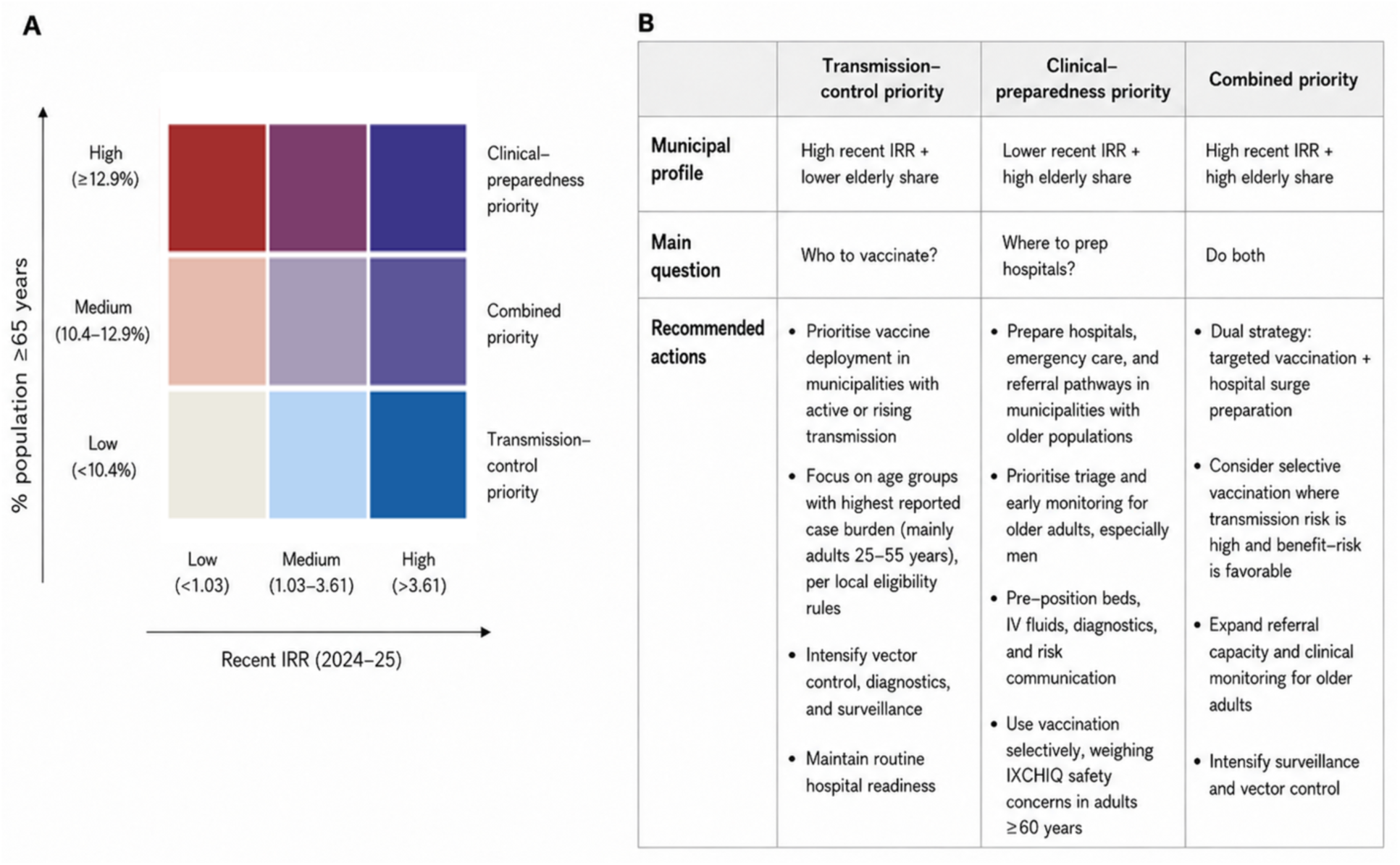
Decision-maker reference for the municipality-level chikungunya vaccination framework, Brazil, 2024–25. (A) Municipality classification using the Figure 6 axes: recent posterior incidence rate ratio (IRR) relative to the national municipality median in 2024–25 (x-axis) and proportion of the municipal population aged ≥65 years (y-axis). Colors correspond to the three priority zones defined by joint empirical tertiles, as in Figure 6. Intermediate cells lean toward the nearest corner action package. (B) Recommended actions by priority zone, organized by municipal profile, the central planning question, and suggested public health responses. Uses the same tertile thresholds as Figure 6: recent IRR <1.03 (low), 1.03–3.61 (medium), >3.61 (high); proportion aged ≥65 years <10.4% (low), 10.4–12.9% (medium), >12.9% (high). This framework is a planning heuristic for SUS implementation; classifications should be updated as surveillance data accumulate and as post-rollout evidence on IXCHIQ safety in adults ≥60 years accrues.

**Figure S9.**
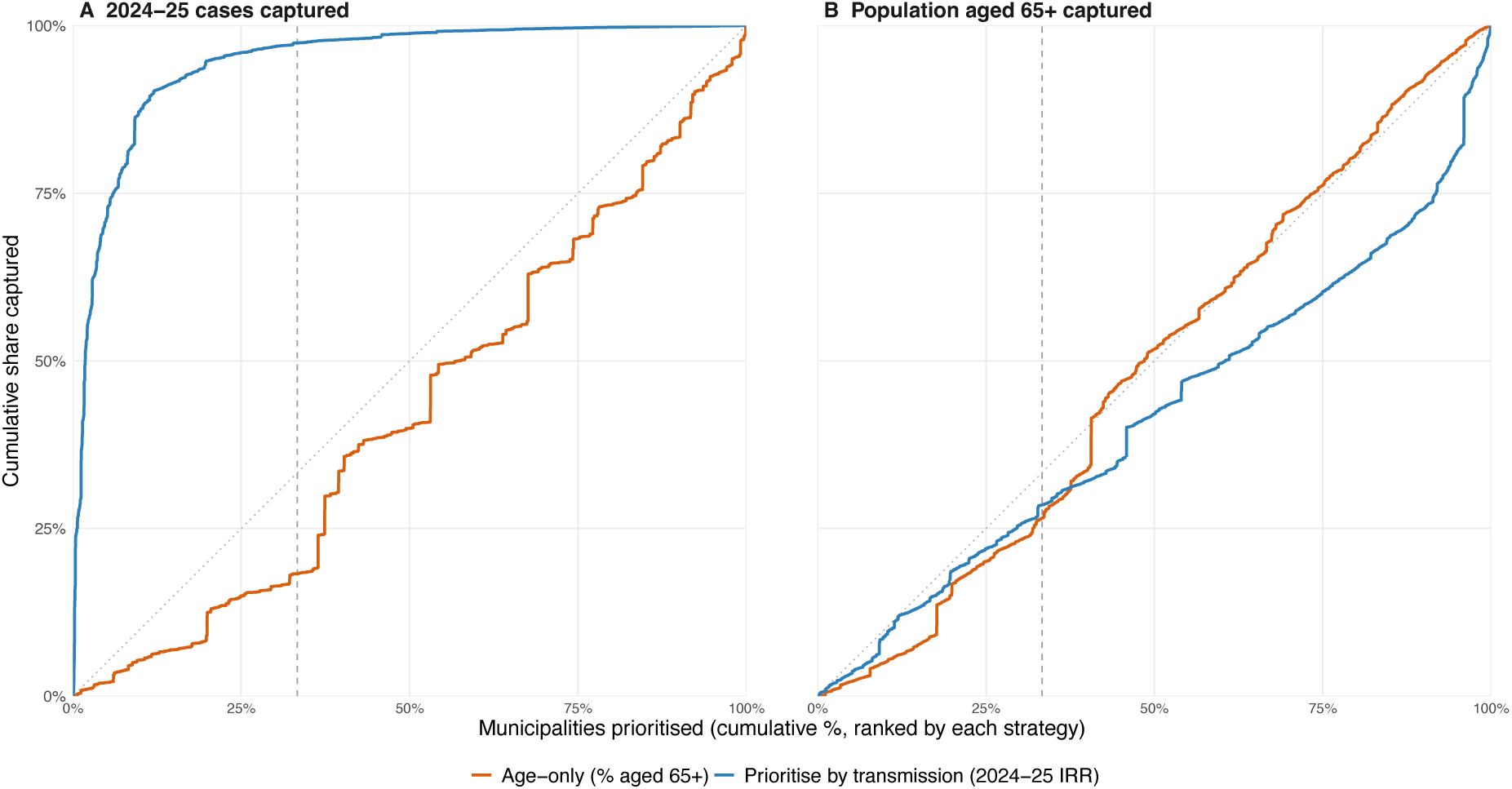
Cumulative capture of 2024–25 reported cases and population aged ≥65 years under two single-axis prioritization strategies, Brazil. (A) Cumulative share of 2024–25 laboratory-confirmed chikungunya cases captured as municipalities are added in descending order of recent posterior IRR (transmission-priority rule, blue) or descending proportion of population aged ≥65 years (age-only rule, orange). (B) Cumulative share of population aged ≥65 years captured under the same two rankings. The dotted diagonal represents random prioritization; the dashed vertical line marks the top tertile threshold (33% of municipalities, n = 1,293). At that threshold, transmission-priority ranking captures 97% of 2024–25 reported cases versus 18% under age-only ranking (Panel A), whereas age-only ranking captures a substantially larger share of the population aged ≥65 years than transmission-priority ranking (Panel B); an age-targeted rule alone therefore leaves 82% of 2024–25 reported cases in municipalities it would not prioritize, which is the gap the transmission axis is designed to close. Neither single axis captures both dimensions efficiently, motivating the joint zoning framework in Figure 6. Municipalities without model estimates (n = 1,691) are excluded. At matched coverage (the 1,293 municipalities constituting the upper tertile of elderly share, ≥12.9% aged ≥65 years), the framework’s clinical-preparedness and combined zones select the same municipalities as an age-only rule and therefore capture an identical share of the severe-outcome-prone population — 26.6% weighted by population aged ≥65 years, and 21.1% weighted by observed 2024–25 deaths among those aged ≥65 years. The framework’s advantage over age-only targeting is thus concentrated on the transmission axis (Panel A), where it recovers the 82% of 2024–25 cases that age-only prioritization would leave uncovered.

### Supplementary Tables

**Table S1.** Published chikungunya seroprevalence studies conducted in Brazil. The table summarizes study locations, survey periods, and target populations extracted from the literature.

| City or area | Survey period (start to end) | Population type | Reference |
| --- | --- | --- | --- |
| Feira de Santana (George Américo) | 2015-11 to 2015-12 | General population (household-based) | <sup>12</sup> |
| Riachão do Jacuípe (Alto do Cemitério) | 2015-11 to 2015-12 | General population (household-based) | <sup>12</sup> |
| Feira de Santana (sentinel areas, urban) | 2016-11 to 2017-09 | General population (household-based, ≥1 year) | <sup>13</sup> |
| Macapá | 2015-01 to 2015-05 | Blood donors (volunteer) | <sup>14</sup> |
| Ribeirão Preto | 2016-01 to 2016-05 | Blood donors (volunteer) | <sup>14</sup> |
| São Sebastião | 2020-02 to 2021-01 | General population (household-based, ≥5 years) | <sup>15</sup> |
| Chapada district, Riachão do Jacuípe | 2016-04 to 2016-04 | General population (household-based, rural residents) | <sup>16</sup> |
| Fulni-ô Indigenous territory (Águas Belas / Itaíba) | 2016-08 to 2017-06 | Indigenous population (Fulni-ô tribe) | <sup>17</sup> |
| Truká Indigenous territory (Cabrobó / Ilha de Assunção) | 2016-08 to 2017-06 | Indigenous population (Truká tribe) | <sup>17</sup> |
| Juazeiro | 2016-08 to 2017-06 | General population (urban control) | <sup>17</sup> |
| Brasília (Federal District) | 2019-01 to 2019-12 | Blood donors (volunteer) | <sup>18</sup> |
| Diamantina | 2018-01 to 2019-12 | Pregnant women (asymptomatic cohort) | <sup>19</sup> |
| Rio de Janeiro city | 2018-07 to 2018-10 | General population (household-based) | <sup>18</sup> |
| Salvador (Pau da Lima community) | 2016-11 to 2017-02 | General population (household-based, ≥5 years) | <sup>19</sup> |
| Salvador | 2017-09 to 2017-11 | Postpartum women (asymptomatic cohort) | <sup>19</sup> |
| Quixadá | 2018-06 to 2019-12 | General population (household-based) | <sup>20</sup> |

**Table S2.** Log-rank tests comparing time-to-event distributions along the chikungunya clinical pathway in Brazil. The table reports log-rank test results for differences in time-to-event distributions stratified by age group, sex, and race or ethnicity across three clinical intervals: symptom onset to hospitalization, hospitalization to death, and symptom onset to death. For each comparison, the number of groups, total sample size, chi-square statistic, degrees of freedom, and p-value are shown.

| Interval | Stratification | $\chi^2$ (df) | p-value |
| --- | --- | --- | --- |
| Onset to hospitalization | Age | 12340 (9) | <2e-16 |
| Onset to hospitalization | Sex | 249 (1) | <2e-16 |
| Onset to hospitalization | Race | 442 (4) | <2e-16 |
| Hospitalization to death | Age | 1107 (9) | <2e-16 |
| Hospitalization to death | Sex | 28.8 (1) | 8.12e-08 |
| Hospitalization to death | Race | 12.6 (4) | 0.0136 |
| Onset to death | Age | 6216 (9) | <2e-16 |
| Onset to death | Sex | 114 (1) | <2e-16 |
| Onset to death | Race | 46.1 (4) | 2.40E-09 |

**Table S3.**
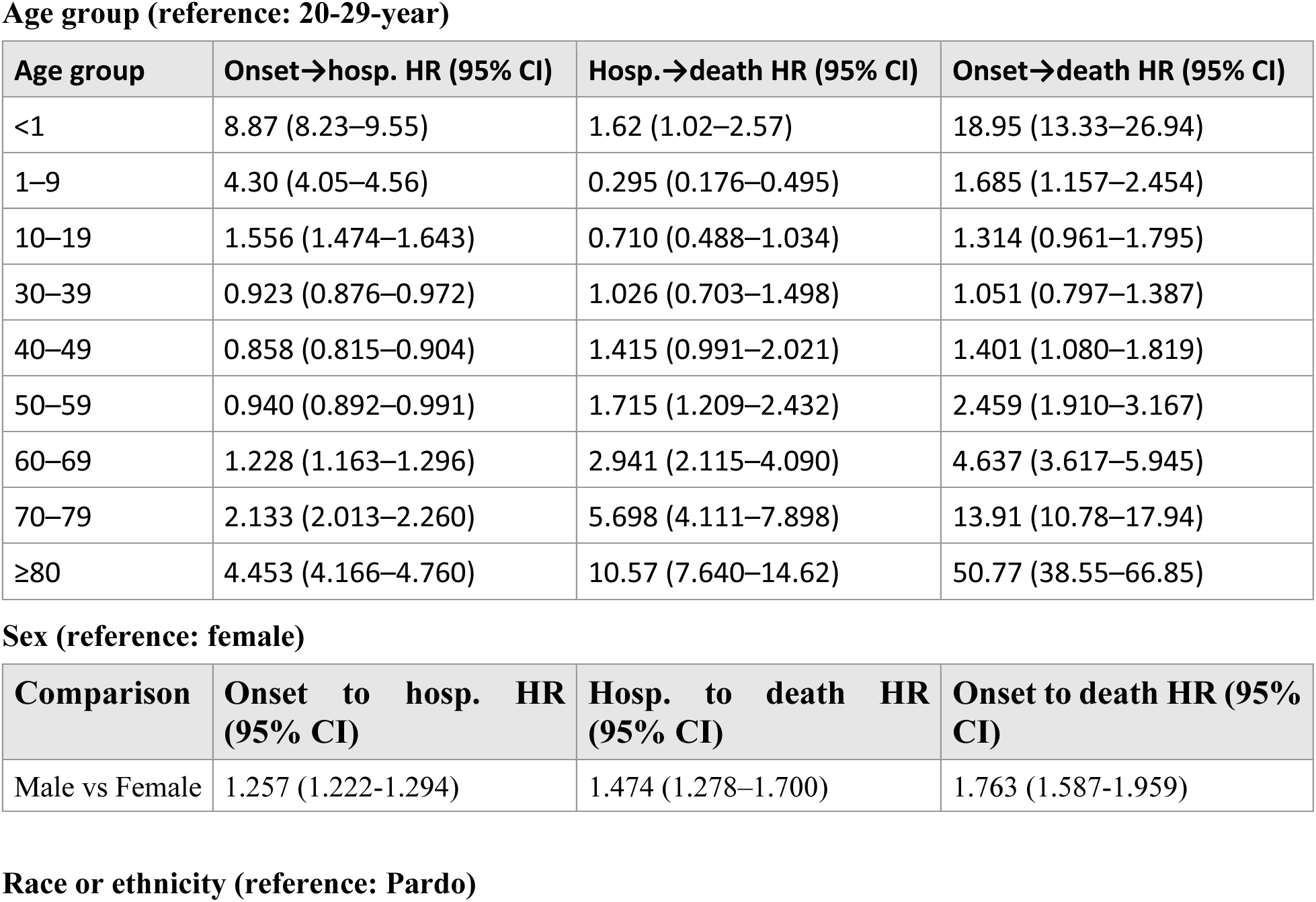
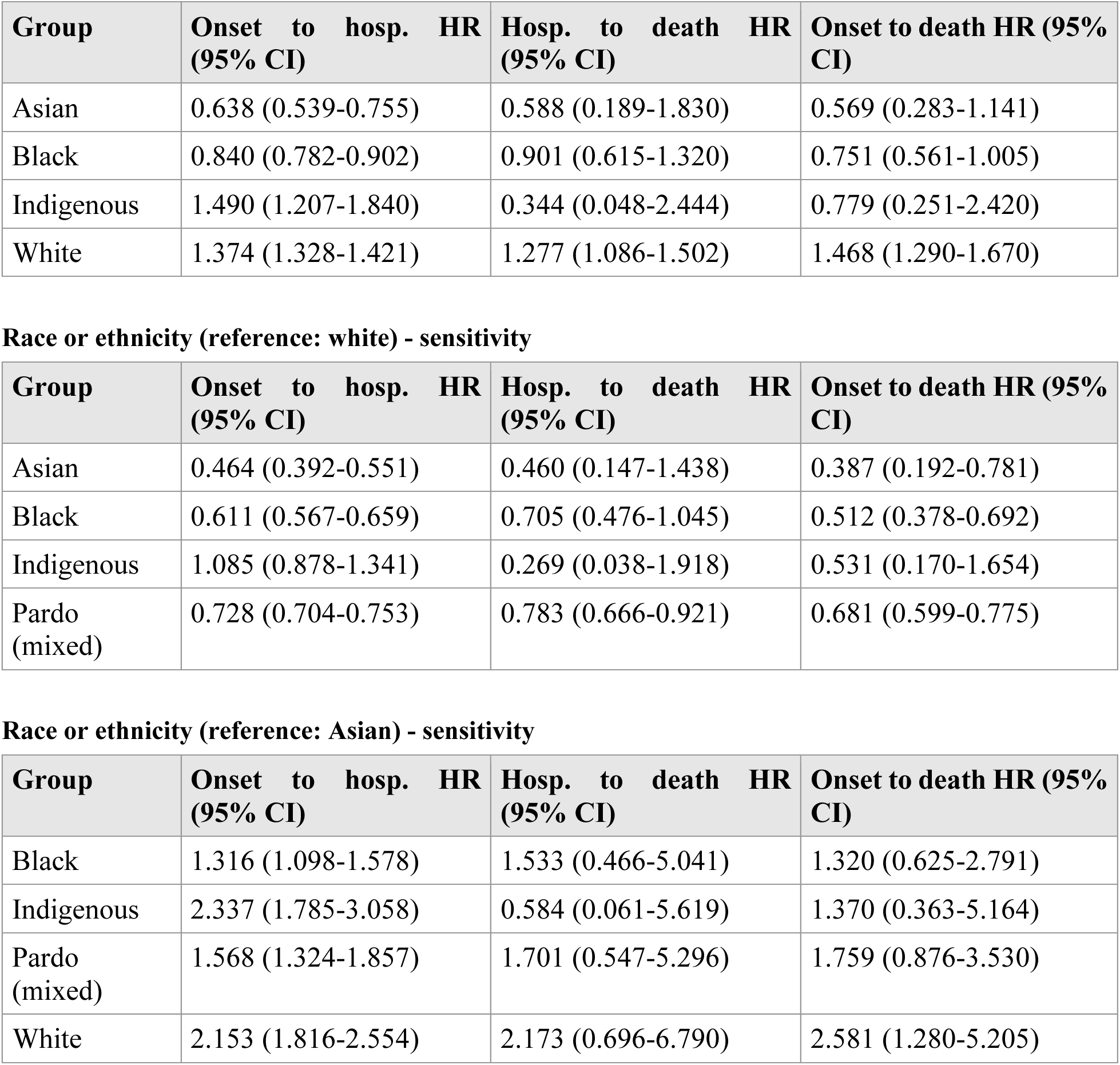
Cox proportional hazards models for demographic predictors of clinical progression among chikungunya cases in Brazil. The table reports hazard ratios (HRs) with 95% confidence intervals (CIs) and p-values for associations of age group, sex, and race or ethnicity with progression across three intervals: symptom onset to hospitalization, hospitalization to death, and symptom onset to death. The race reference group is Pardo (mixed), and age-specific estimates are shown relative to the 20–29 years reference category. Sensitivity analyses use white and Asian as alternative references

**Table S4.**
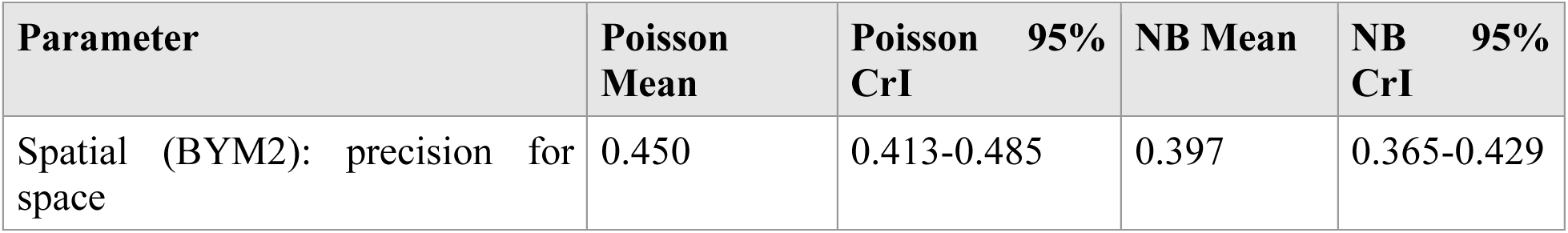
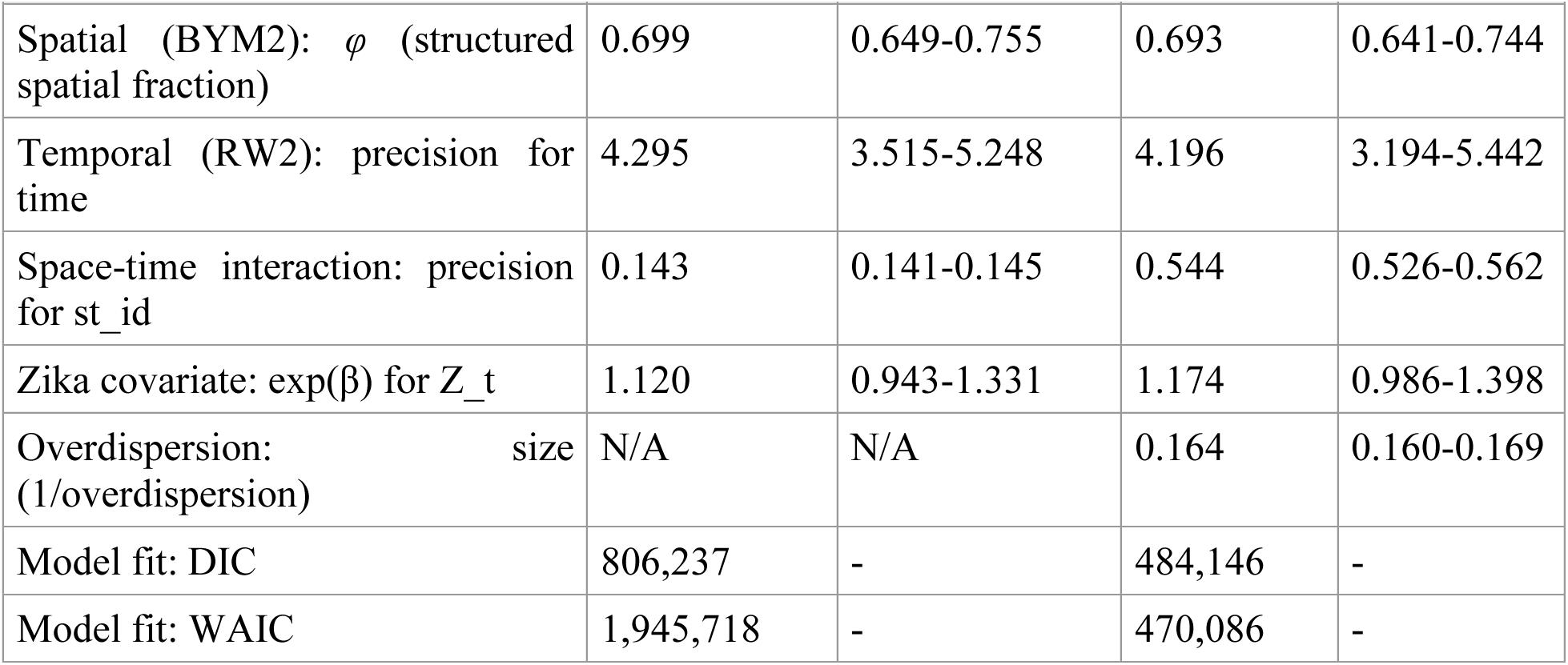
Comparison of key spatiotemporal model parameters between the primary Poisson specification and a negative binomial sensitivity analysis. CrI = credible interval. *φ* = proportion of total spatial variance attributable to structured spatial clustering. The negative binomial model showed improved fit by both DIC and WAIC, confirming overdispersion in municipality-month case counts. Estimates of *φ*, temporal precision, and the Zika covariate association were highly consistent across both specifications, supporting the robustness of the primary model conclusions.

| Parameter | Poisson Mean | Poisson 95% CrI | NB Mean | NB 95% CrI |
| --- | --- | --- | --- | --- |
| Spatial (BYM2): precision for space | 0.450 | 0.413-0.485 | 0.397 | 0.365-0.429 |
| Spatial (BYM2): $\phi$ (structured spatial fraction) | 0.699 | 0.649-0.755 | 0.693 | 0.641-0.744 |
| Temporal (RW2): precision for time | 4.295 | 3.515-5.248 | 4.196 | 3.194-5.442 |
| Space-time interaction: precision for st_id | 0.143 | 0.141-0.145 | 0.544 | 0.526-0.562 |
| Zika covariate: $\exp(\beta)$ for Z_t | 1.120 | 0.943-1.331 | 1.174 | 0.986-1.398 |
| Overdispersion: size (1/overdispersion) | N/A | N/A | 0.164 | 0.160-0.169 |
| Model fit: DIC | 806,237 | - | 484,146 | - |
| Model fit: WAIC | 1,945,718 | - | 470,086 | - |

**Table S5.** Macro-regional case fatality among hospitalized laboratory-confirmed chikungunya cases by epidemic period and relative transmission intensity, Brazil, 2016–17 and 2024–25. Case fatality rate (CFR) was calculated as the number of deaths divided by the number of hospitalized laboratory-confirmed cases within each macro-region and epidemic period. Relative transmission intensity reflects the macro-regional median posterior incidence rate ratio (IRR) from the Bayesian spatiotemporal model (Figure 2), categorized qualitatively as lowest, low, moderate, or highest relative to other macro-regions within the same epidemic period. During the first epidemic wave, the Northeast concentrated both the highest transmission intensity and the highest case fatality (6.36%). By the second wave, this alignment had broken: the Central-West had become the dominant high-transmission region but recorded a moderate case fatality (3.54%), while the Southeast, with moderate transmission intensity, had the highest case fatality (4.95%). Northeast case fatality fell to 1.38% despite the region’s historically severe burden. This progressive spatial discordance between transmission and severity supports the demographic stratification described in the main text.

| Macro-region | Epidemic period | Hospitalized confirmed cases | Deaths | CFR (%) | Relative transmission intensity |
| --- | --- | --- | --- | --- | --- |
| Central-West | Epidemic 1 (2016–17) | 100 | 3 | 3.00 | Low |
| Central-West | Epidemic 2 (2024–25) | 2,285 | 81 | 3.54 | Highest |
| North | Epidemic 1 (2016–17) | 924 | 5 | 0.54 | Moderate |
| North | Epidemic 2 (2024–25) | 329 | 3 | 0.91 | Moderate |
| Northeast | Epidemic 1 (2016–17) | 2,926 | 186 | 6.36 | Highest |
| Northeast | Epidemic 2 (2024–25) | 1,592 | 22 | 1.38 | Moderate |
| South | Epidemic 1 (2016–17) | 17 | 0 | 0.00 | Lowest |
| South | Epidemic 2 (2024–25) | 461 | 14 | 3.04 | Low |
| Southeast | Epidemic 1 (2016–17) | 769 | 17 | 2.21 | Low |
| Southeast | Epidemic 2 (2024–25) | 3,291 | 163 | 4.95 | Moderate |

**Table S6.** Numbers at risk and censored observations for Kaplan-Meier survival analyses along the chikungunya clinical pathway, Brazil, 2015–2025. For each panel, numbers at risk (upper section) and interval-censored observations (lower section) are shown at selected time points for three sequential clinical intervals (symptom onset to hospitalization, hospitalization to death, and symptom onset to death), stratified by age group (Panel A), sex (Panel B), and race or ethnicity (Panel C). Time points are shown in days from the start of each interval. Individuals who did not experience the event of interest were right censored at the last observed date.

| Panel A: Stratification by age group |  |  |  |  |  |  |  |  |  |  |  |  |
| --- | --- | --- | --- | --- | --- | --- | --- | --- | --- | --- | --- | --- |
| Number at Risk |  |  |  |  |  |  |  |  |  |  |  |  |
| Age group | Symptoms onset to hospitalization |  |  |  | Hospitalization to death |  |  |  | Symptoms onset to death |  |  |  |
|  | t=0 | t=50 | t=100 | t=150 | t=0 | t=50 | t=100 | t=150 | t=0 | t=50 | t=100 | t=150 |
| <1 | 10,860 | 9,861 | 9,857 | 9,855 | 1,010 | 341 | 213 | 190 | 10,860 | 10,163 | 10,034 | 10,010 |
| 1–9 | 65,999 | 62,963 | 62,939 | 62,934 | 3,068 | 867 | 470 | 384 | 65,999 | 63,785 | 63,384 | 63,298 |
| 10–19 | 137,999 | 135,692 | 135,662 | 135,655 | 2,356 | 692 | 305 | 233 | 137,999 | 136,312 | 135,924 | 135,852 |
| 20–29 | 200,314 | 198,141 | 198,124 | 198,123 | 2,204 | 567 | 242 | 173 | 200,314 | 198,660 | 198,331 | 198,262 |
| 30–39 | 212,144 | 210,028 | 210,006 | 209,998 | 2,155 | 582 | 226 | 175 | 212,144 | 210,547 | 210,189 | 210,138 |
| 40–49 | 209,744 | 207,799 | 207,776 | 207,773 | 1,983 | 526 | 195 | 138 | 209,744 | 208,255 | 207,918 | 207,861 |
| 50–59 | 178,679 | 176,864 | 176,841 | 176,837 | 1,849 | 504 | 189 | 126 | 178,679 | 177,269 | 176,944 | 176,880 |
| 60–69 | 125,580 | 123,911 | 123,894 | 123,889 | 1,695 | 429 | 150 | 113 | 125,580 | 124,234 | 123,951 | 123,912 |
| 70–79 | 66,348 | 64,820 | 64,809 | 64,807 | 1,548 | 381 | 135 | 97 | 66,348 | 65,067 | 64,815 | 64,773 |

| ≥80 | 26,736 | 25,465 | 25,456 | 25,454 | 1,285 | 263 | 91 | 68 | 26,736 | 25,561 | 25,372 | 25,344 |
| --- | --- | --- | --- | --- | --- | --- | --- | --- | --- | --- | --- | --- |
| Number of Censored |  |  |  |  |  |  |  |  |  |  |  |  |
| Age group | Symptoms onset to hospitalization |  |  |  | Hospitalization to death |  |  |  | Symptoms onset to death |  |  |  |
|  | t=0 | t=50 | t=100 | t=150 | t=0 | t=50 | t=100 | t=150 | t=0 | t=50 | t=100 | t=150 |
| <1 | 0 | 0 | 0 | 0 | 12 | 629 | 124 | 23 | 12 | 629 | 124 | 23 |
| 1–9 | 0 | 0 | 0 | 0 | 32 | 2,173 | 383 | 82 | 32 | 2,173 | 383 | 82 |
| 10–19 | 0 | 0 | 0 | 0 | 19 | 1,625 | 378 | 69 | 19 | 1,625 | 378 | 69 |
| 20–29 | 0 | 0 | 0 | 0 | 35 | 1,587 | 303 | 65 | 35 | 1,587 | 303 | 65 |
| 30–39 | 0 | 0 | 0 | 0 | 53 | 1,495 | 340 | 52 | 53 | 1,495 | 340 | 52 |
| 40–49 | 0 | 0 | 0 | 0 | 35 | 1,382 | 320 | 56 | 35 | 1,382 | 320 | 56 |
| 50–59 | 0 | 0 | 0 | 0 | 31 | 1,272 | 299 | 62 | 31 | 1,272 | 299 | 62 |
| 60–69 | 0 | 0 | 0 | 0 | 22 | 1,167 | 266 | 35 | 22 | 1,167 | 266 | 35 |
| 70–79 | 0 | 0 | 0 | 0 | 19 | 1,016 | 224 | 35 | 19 | 1,016 | 224 | 35 |
| ≥80 | 0 | 0 | 0 | 0 | 15 | 788 | 158 | 22 | 15 | 788 | 158 | 22 |
| Panel B: Stratification by sex |  |  |  |  |  |  |  |  |  |  |  |  |
| Number at Risk |  |  |  |  |  |  |  |  |  |  |  |  |
| Sex | Symptoms onset to hospitalization |  |  |  | Hospitalization to death |  |  |  | Symptoms onset to death |  |  |  |
|  | t=0 | t=50 | t=100 | t=150 | t=0 | t=50 | t=100 | t=150 | t=0 | t=50 | t=100 | t=150 |
| Female | 756,502 | 745,998 | 745,887 | 745,859 | 10,693 | 2,869 | 1,206 | 895 | 756,502 | 748,414 | 746,722 | 746,405 |
| Male | 476,765 | 468,411 | 468,342 | 468,331 | 8,459 | 2,283 | 1,010 | 802 | 476,765 | 470,305 | 469,006 | 468,791 |
| Number of Censored |  |  |  |  |  |  |  |  |  |  |  |  |
| Sex | Symptoms onset to hospitalization |  |  |  | Hospitalization to death |  |  |  | Symptoms onset to death |  |  |  |

|  | t=0 | t=50 | t=100 | t=150 | t=0 | t=50 | t=100 | t=150 | t=0 | t=50 | t=100 | t=150 |
| --- | --- | --- | --- | --- | --- | --- | --- | --- | --- | --- | --- | --- |
| Female | 0 | 0 | 0 | 0 | 169 | 7,390 | 1,585 | 306 | 169 | 7,390 | 1,585 | 306 |
| Male | 0 | 0 | 0 | 0 | 104 | 5,743 | 1,210 | 195 | 104 | 5,743 | 1,210 | 195 |
| Panel C: Stratification by race or ethnicity |  |  |  |  |  |  |  |  |  |  |  |  |
| Number at Risk |  |  |  |  |  |  |  |  |  |  |  |  |
| Race & Ethnicity Group | Symptoms onset to hospitalization |  |  |  | Hospitalization to death |  |  |  | Symptoms onset to death |  |  |  |
|  | t=0 | t=50 | t=100 | t=150 | t=0 | t=50 | t=100 | t=150 | t=0 | t=50 | t=100 | t=150 |
| Pardo (Mixed) | 675,697 | 664,325 | 664,215 | 664,191 | 11,550 | 3,177 | 1,446 | 1,154 | 675,697 | 667,036 | 665,273 | 664,975 |
| Asian | 12,435 | 12,302 | 12,300 | 12,300 | 136 | 30 | 8 | 6 | 12,435 | 12,325 | 12,302 | 12,300 |
| Black | 56,554 | 55,762 | 55,751 | 55,748 | 813 | 230 | 92 | 69 | 56,554 | 55,954 | 55,814 | 55,790 |
| Indigenous | 3,426 | 3,343 | 3,341 | 3,340 | 87 | 14 | 7 | 6 | 3,426 | 3,351 | 3,344 | 3,343 |
| White | 199,586 | 194,978 | 194,939 | 194,931 | 4,672 | 1,164 | 414 | 287 | 199,586 | 195,985 | 195,225 | 195,096 |
| Number of Censored |  |  |  |  |  |  |  |  |  |  |  |  |
| Race & Ethnicity Group | Symptoms onset to hospitalization |  |  |  | Hospitalization to death |  |  |  | Symptoms onset to death |  |  |  |
|  | t=0 | t=50 | t=100 | t=150 | t=0 | t=50 | t=100 | t=150 | t=0 | t=50 | t=100 | t=150 |
| Pardo (Mixed) | 0 | 0 | 0 | 0 | 157 | 7,886 | 1,643 | 282 | 157 | 7,886 | 1,643 | 282 |
| Asian | 0 | 0 | 0 | 0 | 2 | 103 | 20 | 2 | 2 | 103 | 20 | 2 |
| Black | 0 | 0 | 0 | 0 | 7 | 553 | 133 | 23 | 7 | 553 | 133 | 23 |
| Indigenous | 0 | 0 | 0 | 0 | 3 | 69 | 7 | 1 | 3 | 69 | 7 | 1 |
| White | 0 | 0 | 0 | 0 | 65 | 3,259 | 721 | 121 | 65 | 3,259 | 721 | 121 |

**Table S7.** Macro-regional distribution of priority zones from the combined geographic and demographic targeting framework, Brazil, 2024 to 2025. Counts of Brazilian municipalities falling into each priority zone (transmission-control, clinical-preparedness, combined), stratified by macro-region. Zones are defined by joint empirical tertiles of recent (2024 to 2025) posterior incidence rate ratio and proportion of population aged 65 years and older (Figure 6). Equal column totals (832 transmission-control, 832 clinical-preparedness) are a mechanical consequence of the tertile classification rather than an empirical finding; macro-regional asymmetry within columns reflects the empirical patterns described in the main text.

| Macro-region | Transmission control | Clinical preparedness | Both |
| --- | --- | --- | --- |
| North | 63 | 9 | 3 |
| Northeast | 367 | 224 | 115 |
| Central-West | 208 | 33 | 39 |
| Southeast | 175 | 395 | 272 |
| South | 19 | 171 | 32 |
| **Total** | **832** | **832** | **461** |

